# Enhancing Cognitive Restructuring with Concurrent Repetitive Transcranial Magnetic Stimulation: A Transdiagnostic Randomized Controlled Trial

**DOI:** 10.1101/2021.01.18.21250060

**Authors:** Andrada D. Neacsiu, Lysianne Beynel, John P. Powers, Steven T. Szabo, Lawrence G. Appelbaum, Sarah H. Lisanby, Kevin S. LaBar

**Affiliations:** Duke University Medical Center; Duke University; Durham VAMC; National Institutes of Health

**Keywords:** cognitive control, transdiagnostic, emotional dysregulation, rTMS, intervention

## Abstract

**Introduction:** Emotional dysregulation constitutes a serious public health problem in need of novel transdiagnostic treatments.

**Objective:** To this aim, we developed and tested a one-time intervention that integrates behavioral skills training with concurrent repetitive transcranial magnetic stimulation (rTMS).

**Methods:** Forty-six adults who met criteria for at least one DSM-5 disorder and self-reported low use of cognitive restructuring (CR) were enrolled in a randomized, double-blind, sham-controlled trial that used a between-subjects design. Participants were taught CR and underwent active rTMS applied at 10 Hz over the right (n= 17) or left (n= 14) dorsolateral prefrontal cortex (dlPFC) or sham rTMS (n= 15) while practicing reframing and emotional distancing in response to autobiographical stressors.

**Results:** Those who received active left or active right as opposed to sham rTMS exhibited enhanced regulation (*d*s = 0.21 - 0.62) as measured by psychophysiological indices during the intervention (higher high-frequency heart rate variability, lower regulation duration). Those who received active rTMS over the left DLPFC also self-reported reduced distress througout the intervention (*d* = 0.30), higher likelihood to use CR, and lower daily distress during the week following the intervention. The procedures were acceptable and feasible with few side effects.

**Conclusions:** These findings show that engaging frontal circuits simultaneously with cognitive skills training and rTMS may be clinically feasible, well-tolerated and may show promise for the treatment of transdiagnostic emotional dysregulation. Larger follow up studies are needed to confirm the efficacy of this novel therapeutic approach.

## Introduction

Emotional dysregulation, defined as a deficit in the ability to reduce or change negative emotional states[1], occurs across several forms of psychopathology [2,3] and drives the severity and duration of a variety of mental health problems[4,5]. Difficulties in emotion regulation can be treated through psychotherapy[2,6-8] in transdiagnostic individuals[9,10]. Transdiagnostic approaches teach emotion regulation skills such as cognitive restructuring (CR), relaxation, and mindfulness[11-14]. However, psychotherapy is considered to be moderately effective in the treatment of emotion dysregulation[9,15] and is still in need for improvement to maximize gains and reduce treatment burden[16]. Specifically, more work is needed to accelerate the process of learning within therapy sessions, to retain skills after therapy sessions, and to help patients generalize in-session learning to out-of-session contexts[17].

One approach to optimize transdiagnostic interventions is to translate findings from basic neuroscience studies on emotion regulation into the development of new interventions. Emerging neuroscientific findings highlight that across several disorders, hypoactivation in prefrontal regions such as the dorsolateral (dlPFC), ventrolateral (vlPFC), and medial (mPFC) prefrontal cortex, and hyperactivation in limbic/paralimbic brain areas, such as the left anterior insula[18,19], characterize difficulties in changing emotional arousal. Brain stimulation can alter pathological brain circuits resulting in translational potential to mitigate problems with emotion regulation. Repetitive transcranial magnetic stimulation (rTMS)[20], in particular, is one type of noninvasive brain stimulation leads to downstream effects through network connectivity, thereby modulating broad brain circuits that underlie psychopathologies [21]. Given the critical role of the dlPFC in the emotion regulation neural network, applying high frequency rTMS --known to have facilitatory effects--over the dlPFC could have potential to regulate its hypoactivation, enhance synaptic plasticity[22], and, as a result, improve learning of emotion regulation skills. It has been demonstrated that excitatory stimulation of the left dlPFC facilitates disengagement from angry faces[23], while excitatory stimulation of the right dlPFC enhances cognitively-based emotion regulation in healthy adults[24].

Concurrent neurostimulation can be used as an augmentation tool for behavioral interventions, with effects apparent even after one session. For example, rTMS can remediate hypoactive brain circuits by enhancing spontaneous activity in targeted regions (e.g., remediating working memory performance following sleep deprivation[25]). In addition, when combined with a behavioral prime (e.g., food, drug paraphernalia, cues that trigger obsessions), active neurostimulation reduces problematic behaviors (respectively, binge eating, drug use, compulsive behaviors) above and beyond sham stimulation[26-28] in the short term. Past studies have combined rTMS with 16-20 sessions of psychotherapy, demonstrating feasibility[29], with enhanced effects over psychotherapy alone[30] or over cognitive training alone[31]. Taken together, these studies show promise for neurostimulation as an augmentation tool to alter emotion regulation.

This randomized, double-blind, sham-controlled study, using a parallel design, aimed to establish the feasibility, acceptability and preliminary efficacy of enhancing emotion regulation skills training for transdiagnostic clinical adults with excitatory rTMS over the dlPFC, concurrently during an emotion regulation task. We selected cognitive restructuring (CR) as a targeted emotion regulation skill – that involves thinking differently about a situation to reduce negative affect[32,33] – because it is generally effective at downregulating emotions[33,34]; it can be implemented across a broad range of affective contexts[34]; and it engages the same fronto-limbic neural networks on which neurostimulation has already been tested[35,36]. To establish proof-of-concept and feasibility, we limited the intervention to a single session and collected high frequency heart rate variability (HF-HRV) as an objective physiological measure of emotion regulation[37-42]. Using a potentiation design[43] we hypothesized that receiving active versus sham rTMS in conjunction with CR would lead to faster recovery from emotional distress during the intervention session, reduced arousal and increased use of CR during the week after the intervention, and improved emotional dysregulation, psychopathology and functioning for up to a month following the intervention.

## Materials and Methods

### Participants and Procedures

This study (N = 83) and an adjunctive supplement (N = 65) were pre-registered together (NCT02573246), and they ran concurrently from April 2016 to February 2020. The results from the supplemental study are not included here. We expected moderate to large effect size differences between regulation during active and sham conditions in our psychophisiological measurements based on previous findings[24]. A G*Power 3.1[44] power analysis indicated that a total sample size of forty-two participants (14/condition) in a repeated measures ANOVA (5-measurements) examining between factors effects, with high correlation between repeated measurements (*r* = 0.70)[45] would be needed to observe the a moderate to large effect size (*f*= .45) with an observed power of .80 and an alpha of .05.

Out of 83 participants enrolled, forty-seven (8 men; 39 women, mean_age_: 30.02 ± 10.73) qualified and were randomized to one of three treatment conditions. The first randomized participant piloted all procedures, and the remaining 46 were considered intent to treat (ITT). Participants self-reported below-average use of CR when upset, were transdiagnostic and met criteria for an average of 2.53 (*SD =* 1.65) current and 4.20 (*SD* = 1.94) lifetime DSM-5 diagnoses (SCID-5)[46]. A third of our sample (31.92%) met criteria for at least one personality disorder (SCID-5-PD)[47]. The CONSORT flow diagram, Figure 2A and the Supplemental Materials detail study flow and inclusion/exclusion criteria. The study was approved by the Duke University Health System (DUHS) Institutional Review Board, and participants were paid for the study visits that they attended. Intake Session Qualified participants completed a clinical interview, a battery of self-reports, and a standardized assessment [48,49] which yielded four autobiographical negative emotional arousal scripts (see Supplemental Materials). Participants were then randomly assigned to active right dlPFC (*n* = 17), active left dlPFC (*n* = 14), or sham stimulation (*n* = 15), matching for biological sex and use of psychotropic medication using a minimization algorithm [50,51] and a 1:1:1 ratio. Sham participants were further randomized using a 1:1 ratio to receive TMS over either the right (*n* = 8) or left DLPFC (*n* = 7).

**Fig. 1.**
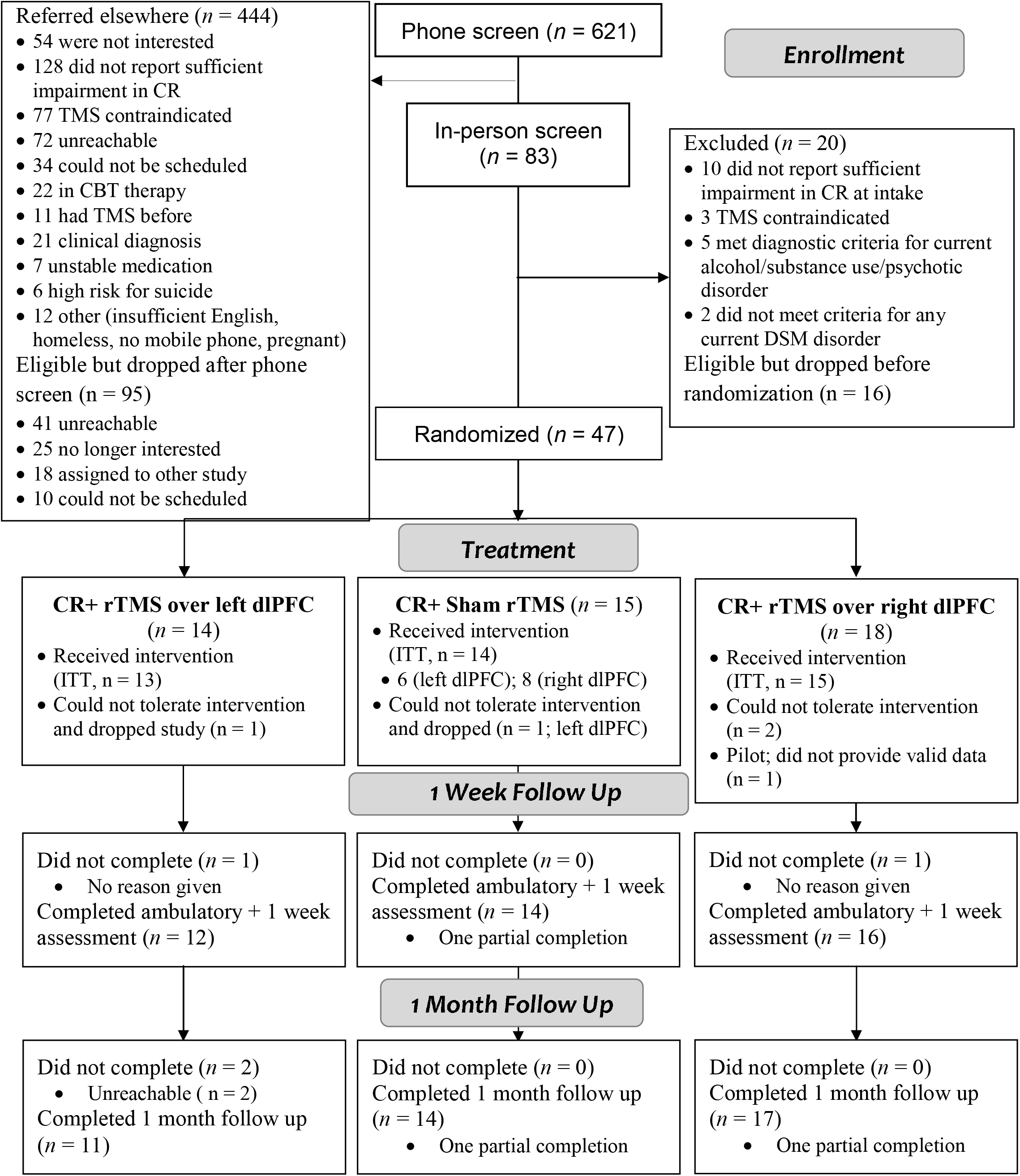
CONSORT Flow Diagram. CR = Cognitive restructuring; rTMS = repetitive transcranial magnetic stimulation; ITT = Intent to Treat; dlPFC = dorsolateral prefrontal cortex

**Fig. 2.**
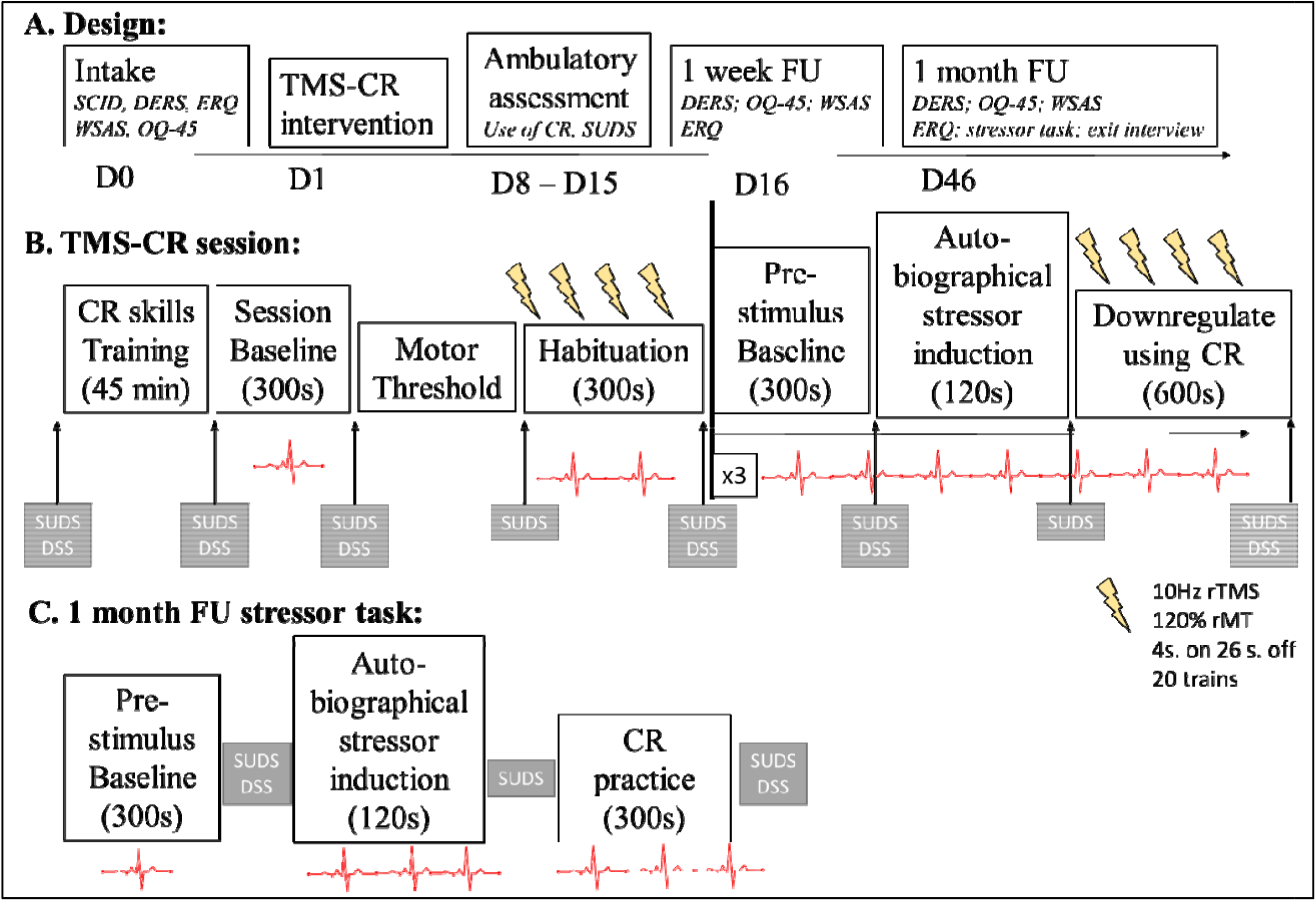
A. Overall study design (with target days since intake when each session was planned to occur), B. Schematic of the TMS-CR intervention. Physiological data were collected during a 300 s rest period (session baseline), followed by the identification of the resting motor threshold (rMT). A 600 s habituation period followed when participants received active or sham rTMS alone while listening to white noise and being instructed to think of nothing in particular. Next, participants were instructed to sit quietly for a 300 s pre-stimulus baseline, followed by instructions to imagine as vividly as possible a negative experience while hearing a 40-s recording of the autobiographical stressor (120 s total), and by instructions to reduce distress using CR (600 s). The rTMS began right after the CR instruction audio was played and continued throughout the regulation period. Every 180s there were prompts to either distance or reframe thoughts related to the autobiographical stressor. Afterwards, there was a break, followed by a second and third administration of the stressor task using the procedures outlined above, but with different personalized stressors presented in a randomized order. C. Schematic of the 1 month follow up stressor task, TMS = repetitive transcranial magnetic stimulation (either active or sham); CR = cognitive restructuring; rTM = resting motor threshold; SUDS = subjective units of distress; DSS = dissociation; SCID = structured clinical interview; DERS = Difficulties in Emotion Regulation; QQ-45 = Outcome Questionnaire – 45; WSAS = Work and social adjustment scale; ERQ = Emotion regulation questionnaire

### Intervention Session

Participants returned for the 3.5-hour intervention session within a month of intake (Fig. 2B). The session started with one-on-one skills training with a trained psychologist, and was focused on learning and practicing CR. Skills training used established procedures blending psychotherapeutic approaches [52,53] with instructions in CR that matched prior neuroimaging studies [54] and included non-specific factors such as unconditional positive regard, attention from the experimenter, and openenss for disclosure from the participant[43]. Both distancing and reframing CR tactics were presented and practiced (see Supplemental Materials).

Before and after the intervention session, participants rated on a scale from 0 to 3 (absent -severe) the intensity of their headache, neck and scalp pain, and hearing impairment. Throughout the intervention day participants rated subjective units of distress (SUDS; 0 to 9 scale) and dissociation [55]. Psychophysiological measurements were collected continuously using the BIOPAC MP-150 recording system (Goleta, CA). The progression of the combined intervention is detailed in Fig. 2B.

### rTMS Parameters

Active or sham rTMS was performed with a figure-8 coil (A/P Cool-B65) and a MagPro X100 stimulator with MagOption (MagVenture, Denmark). Stimulation was delivered over the left or right dlPFC, defined according to the 10-20 system [56](Supplementary Fig.1). A stereotaxic neuronavigation system was used (Brainsight, Rogue Research), and a template brain (MNI) was registered to each participant’s head using anatomical landmarks, to maintain accurate coil positioning across the session. Twenty trains of 10-Hz rTMS (4 s, 26 s inter-train interval) were delivered at 120% of rMT [57-59] four times over the course of the intervention session for a total of 3200 pulses. Sham stimulation was delivered using the opposite, shielded face of the same A/P coil. Sham electrodes delivering a weak electrical current on the scalp [60], in order to mimic the active TMS-induced somato-sensory sensations were put near the hairline for all participants and were only activated for sham participants.

### Ambulatory Assessment and Follow-Up Visit

Following the intervention participants received 8 calls/day for 7 days, starting the day after the intervention, at pseudo-random times. On each call participants were asked if they had used CR since the previous call (yes/no) and their current level of distress (0 to 9). At the end of the week, the battery of self-reports from intake was administered again via an online link.

Participants returned to the research office a month later to complete a stressor task with a 4^th^ autobiographical stressor and without rTMS (Fig. 2C), an exit interview, and self-reports. Afterwards, the blind was broken and the experimenter and the subject debriefed.

### Measures

The primary outcomes for the study were HF-HRV during emotion regulation and the time it took to downregulate autobiographically induced distress. During the intervention and follow-up sessions, each period (e.g., baseline, habituation, stressor, and regulation) was divided into 120-s bins, and HF-HRV was extracted from the cleaned ECG signal from each bin following established guidelines [61]. For each baseline, heart rate (HR) was averaged from the last 240 s of the total 300 s, excluding any disruptions. Time to return to one’s HR baseline during the regulation period (regulation duration) was defined as the amount of time it took from the beginning of regulation for the continuously monitored HR to reach a value that was lower or equal to the average pre-stimulus baseline HR. A baseline value for regulation duration was computed as the time it took during habituation for the person to return to HR baseline after the rTMS driven increased arousal.

Secondary outcomes included acceptability and feasibility metrics, self-reports related to psychopathology and emotional dysregulation (ERQ, Difficulties in Emotion Regulation Scale --DERS[62], Outcome Questionnaire --OQ-45[63], Work and Social Adjustment Scale --WSAS[64]), SUDS and use of CR during the ambulatory week. Participants were asked to give their best guess whether they received real or sham rTMS (forced-choice question) and to rate their confidence in their response after the intervention and at the 1-week and 1-month follow-up assessments. An exit interview [29] was administered at 1-month to examine perceived feasibility and acceptability on a scale from 0 (not at all) to 9 (extremely). Satisfaction was rated on a 0 (low) to 100 (high) scale.

### Statistical Analyses

Mixed-effects hierarchical linear models (MMANOVA) with analytically determined covariance structures were used to analyze the repeated measures data [65] (see Supplemental Materials for the SPSS syntax for the models used). Estimated marginal means (EMMs) were compared using LSD corrections for significant main and interaction effects. Effect sizes for these models were computed using Feingold’s formula [66] and interpreted using Cohen’s [67] specifications. To test immediate effects of the intervention, we conducted three analyses examining HF-HRV, regulation duration, and SUDS using a Benjamini and Hochberg false discovery rate (FDR) correction [68](*q* = 0.10). See Supplemental Materials for details. Generalized estimated equations models (GEE) [69] using ordinal logistic models and an independent covariance structure examined differences between treatment conditions in side effects.

To test near-term effects of the TMS-CR intervention, a hierarchical linear model (HLM) [70] was used to examine condition differences in SUDS and a GEE binary logistic model (logit link, unstructured working correlation with robust estimators) [71,72] examined differences in use of CR (indicated as yes/no on each call) during the ambulatory week. An FDR correction [68] was employed for these analyses (*q* = 0.10).

To test the long-term effects of the intervention, six MMANOVA models were conducted: four examining between-condition differences at the 1-week and 1-month follow-up assessments in ERQ, DERS, OQ-45, and WSAS; and two examining HF-HRV and SUDS during the follow-up stressor task. An ANCOVA examined differences in regulation duration at follow up. The significance threshold for the seven long-term analyses was adjusted using an FDR correction [68]. In cases where significant differences were not found, equivalence testing was employed using the two one-sided test procedure[73,74] with symmetric equivalence margins of .5 as suggested by others [75,76].

## Results

### Missing Data

Out of 46 ITT participants, four could not tolerate the target rTMS dose; nevertheless, all available data are included in the final outcome analyses (see Table 1 for demographics). Across the variables examined for those included, only 4.3% of the data was missing (Supplemental Materials).

**Table 1.** Demographics and Clinical Descriptives by Group

|  | Active Left (*n* = 14) | Active Right (*n* = 17) | Sham (*n* = 15) |
| --- | --- | --- | --- |
| Mean age (SD) | 33.29 (13.98) | 27.76 (7.23) | 29.53 (10.56) |
| Female gender (%) | 78.6 | 82.4 | 86.7 |
| Latinx Background (%) | 21.4 | 11.8 | 6.7 |
| Racial Background (%) |  |  |  |
| Caucasian | 78.6 | 52.9 | 66.7 |
| African American | 14.4 | 11.8 | 13.3 |
| Asian American | 7.10 | 23.6 | 20.1 |
| More than one racial group | 0.00 | 11.8 | 0.00 |
| Sexual Orientation (%) |  |  |  |
| Heterosexual | 78.6 | 56.3 | 73.3 |
| Homosexual | 14.3 | 11.8 | 0 |
| Bisexual | 7.1 | 29.4 | 13.3 |
| Currently in relationship (%) |  |  |  |
| Single never married | 64.3 | 70.6 | 60.0 |
| Married | 14.3 | 17.6 | 26.7 |
| Divorced/separated | 7.1 | 0.00 | 6.7 |
| Living with Partner | 14.3 | 11.8 | 6.7 |
| Household Income (%) |  |  |  |
| \$0-\$10,000 | 14.3 | 17.6 | 13.3 |
| \$10,001 - \$20,000 | 7.1 | 17.6 | 0.00 |
| \$20,000 - \$40,000 | 28.6 | 23.5 | 20.0 |
| \$40,001 - \$65,000 | 14.3 | 11.8 | 20.0 |
| \$65,001 - \$100,000 | 14.3 | 11.8 | 13.3 |
| more than \$100,000 | 21.4 | 17.6 | 33.3 |
| Total # of diagnoses current | 2.86 (1.56) | 2.24 (1.64) | 2.53 (1.85) |
| Total # of diagnoses in lifetime | 4.71 (1.73) | 3.88 (1.80) | 3.80 (2.08) |
| Current Disorders (%) |  |  |  |
| Depressive Disorders | 50.0 | 41.2 | 26.7 |
| Bipolar Disorder | 7.1 | 5.9 | 0.00 |
| Anxiety Disorders | 85.7 | 76.5 | 73.3 |
| Lifetime Disorders (%) |  |  |  |
| Depressive Disorders | 92.9 | 70.6 | 86.7 |
| Anxiety Disorders | 92.9 | 82.4 | 73.3 |
| Substance Use | 7.1 | 17.6 | 6.7 |
| Avoidant PD (%) | 7.1 | 11.8 | 6.7 |
| Borderline PD (%) | 14.3 | 0.0 | 0.0 |
| Paranoid PD (%) | 7.1 | 0.0 | 0.0 |
| Histrionic PD (%) | 0.00 | 0.00 | 6.7 |
| Obsessive-Compulsive PD (%) | 42.9 | 0.00 | 20.0 |
| Any PD (%) | 50.0 | 17.6 | 26.7 |
| Taking psychotropic meds (%) | 14.3 | 17.6 | 26.7 |

*Note.* PD = personality disorder

### Normality Assumption

At both intervention and follow up, HF-HRV was transformed using the function lg10*(HF-HRV*1000000) and regulation duration was transformed with a lg10 function. Intervention SUDS were also transformed to a normal distribution using a square root function.

### Tolerability and Acceptability

Participants found the overall study procedures very acceptable (M_Acceptability_ = 7.26, SD = 1.08) and feasible (M_Feasibility_ = 6.61, SD = 0.79) with no differences between conditions (*p*s > .05). Participants were 59.40% (SD = 33.51) likely to recommend this treatment to a friend. Participants reported low distress induced by neurostimulation while they were engaging in CR (M = 2.19; SD = 1.04) and moderate-to-low interference with concentrating from the TMS noise (M= 4.80, SD = 2.47). When asked the open-ended question “what was it like being in our study?”, all participants reported that they had a positive experience, found CR very useful, and thought it had a positive impact on their lives. There were no serious adverse events in the study.

There was no significant difference between treatment conditions in headache (Wald χ^2^[2] = 0.11, *p* = .95) and neck pain (Wald χ^2^[2] = 2.61, *p* = .27). Five participants in the active left condition (35.71%), eight in the active right condition (47.05%), and six in the sham condition (40.0%) reported worsened headache after the intervention. One participant in the active left condition reported mild hearing problems following our procedures. One participant from the active left and four from the active right condition reported mild scalp discomfort following the intervention. No one experienced a seizure, and all side effects resolved by the next study contact.

All participants reported being naïve to rTMS at the beginning of treatment. Three participants in the active conditions and five in the sham conditions believed they received sham rTMS when asked to guess their condition assignment (χ^2^ [2] = 5.68, *p* = .06). Nevertheless, the majority of participants were not confident in their choice (53.5% indicated it could have been either) or thought they more likely than not received real stimulation (47.4%). All participants who guessed sham rated their confidence in their assignment as a 5 (uncertain).There was no significant difference between conditions in confidence about assigned condition right after the intervention (*F*[2,42] = 1.92, *p* = .16), or at the one week (*F*[2,38] = 1.15, *p* = .33), or one month follow up (*F*[2,41] = 0.15, *p* = .86). These findings suggest that the procedures are feasible and acceptable and that the blinding procedures described were successful.

### The Effect of the Combined Intervention on Immediate Emotion Regulation

To test whether neurostimulation would enhance emotion regulation at the moment when it is administered, we conducted three MMANOVA analyses using HF-HRV during regulation, SUDS after regulation, and regulation duration as dependent variables.

#### Heart Rate Variability Results

The MMANOVA analysis of HF-HRV, using an unstructured covariance, showed a significant main effect of treatment condition (*F*[2, 32.17] = 4.34, *p* =.02, *p*_*FDR*_ = .03), time within each regulation period (*F*[4, 39.19] = 6.17, *p* <.005), session baseline (*F*[1, 66.70] = 104.27), and pre-stimulus baseline *F*[1, 81.25] = 59.39, *p* <.001), as well as a significant treatment condition by time interaction (*F*[8, 39.24] = 2.77, *p* < .05). Active rTMS over the left (EMM_*lg_HF-HRV*_ = 1.91, S.E.= .04) and right dlPFC (EMM_*lg_HF-HRV*_= 1.86, *S*.*E*. = .04) led to significantly higher HF-HRV (i.e., enhanced emotion regulation) when compared to sham rTMS (EMM_*lg_HF-HRV*_= 1.75, *S*.*E*. = .04), *p*s < .05 (*d*s = 0.21 -0.31). The active rTMS conditions were not significantly different (*p =* .34) and were equivalent (90% CI = [-0.38 -0.46]). LSD-corrected pairwise comparisons of the interaction effect showed a significant difference between active and sham stimulation for the first 6 minutes of regulation (*p*s < .05), and no difference for the last 4 minutes, suggesting that sham participants “caught up” with active participants in their emotion regulation by the end of the regulation period (See Fig. 3). The effect size of the difference between sham and active TMS was largest (Cohen’s *d* =.45-.53) in the first 2 minutes of regulation.

**Fig. 3.**
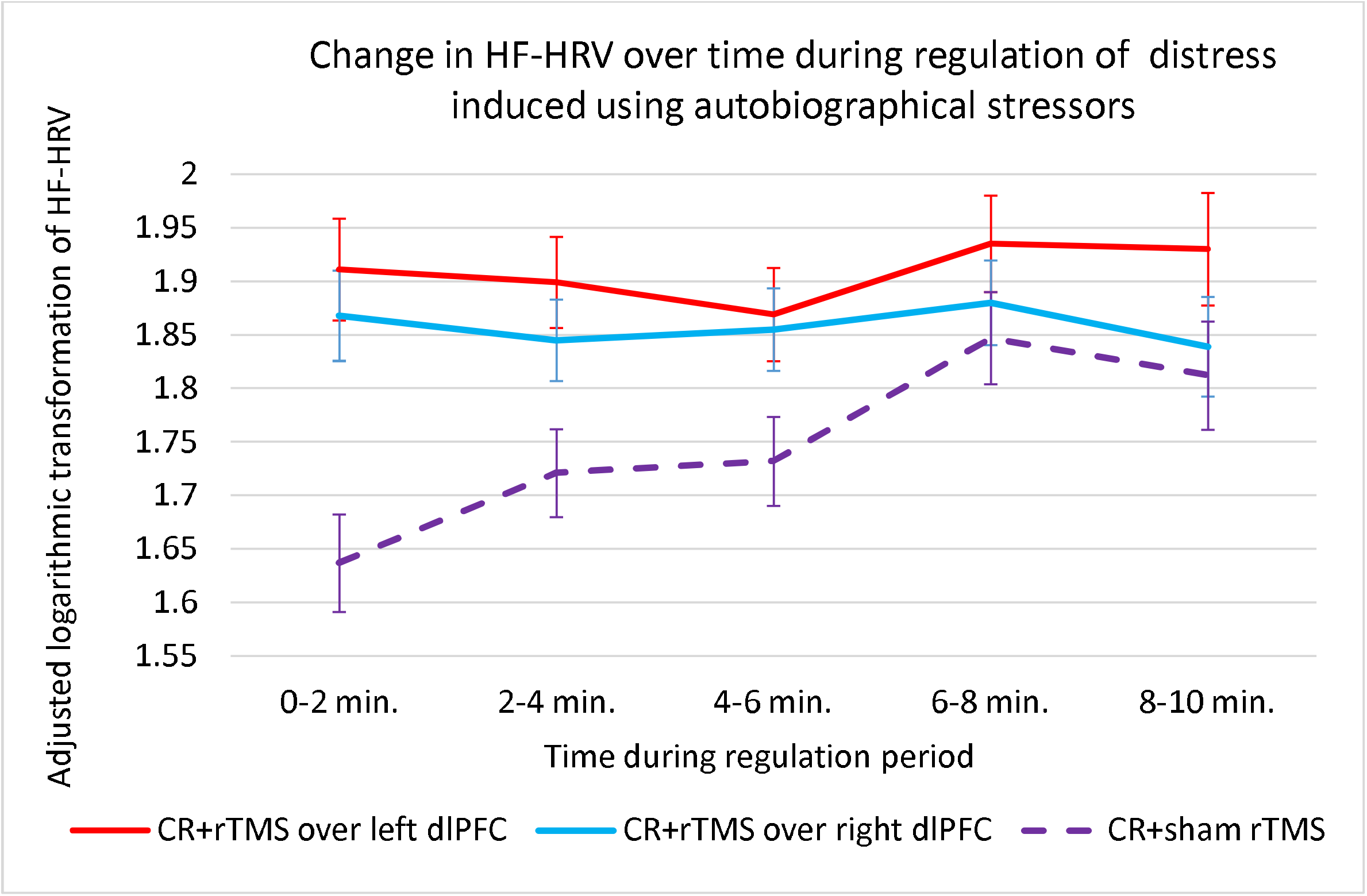
Changes in high frequency heart rate variability (HF-HRV; a marker of effective emotional regulation) summarized across the three regulation periods allotted during the intervention, and separated by condition, and by time segment. The original HF-HRV value was measured in seconds^2^, multiplied by 100000 and transformed using a log function to achieve normality. Each time point on the graph represents the estimate marginal mean from the LSD pairwise comparisons from the MMANOVA main analysis which accounts for covariates. Regulation periods followed autobiographical negative emotional inductions.

#### Self-Report Results

The experimental instructions (i.e., try to relax, remember a stressor, use CR) successfully modulated distress across conditions (*F*[2, 40.32] = 70.97, *p* < .001; Fig. 4). Participants in the active left rTMS condition experienced significantly lower distress (EMM_SR_SUDS_ = 1.19; S.E. = .08) during the intervention when compared to sham (EMM_SR_SUDS_ = 1.43; S.E. = .08; *d* = .30; *p* < .05) and active right rTMS participants (EMM_SR_SUDS_ = 1.44; S.E. = .08, *d* = .32; *p* < .05; *F*_*treatment_condition*_[2,34.99] = 3.41, *p* = .04, *p*_*FDR*_ = .06). LSD-corrected pairwise comparisons did not find a significant difference between right and sham rTMS. The treatment condition by instruction interaction was not significant (*F*[4, 40.62] = 1.88, *p* = .13; see Fig. 4).

**Fig. 4.**
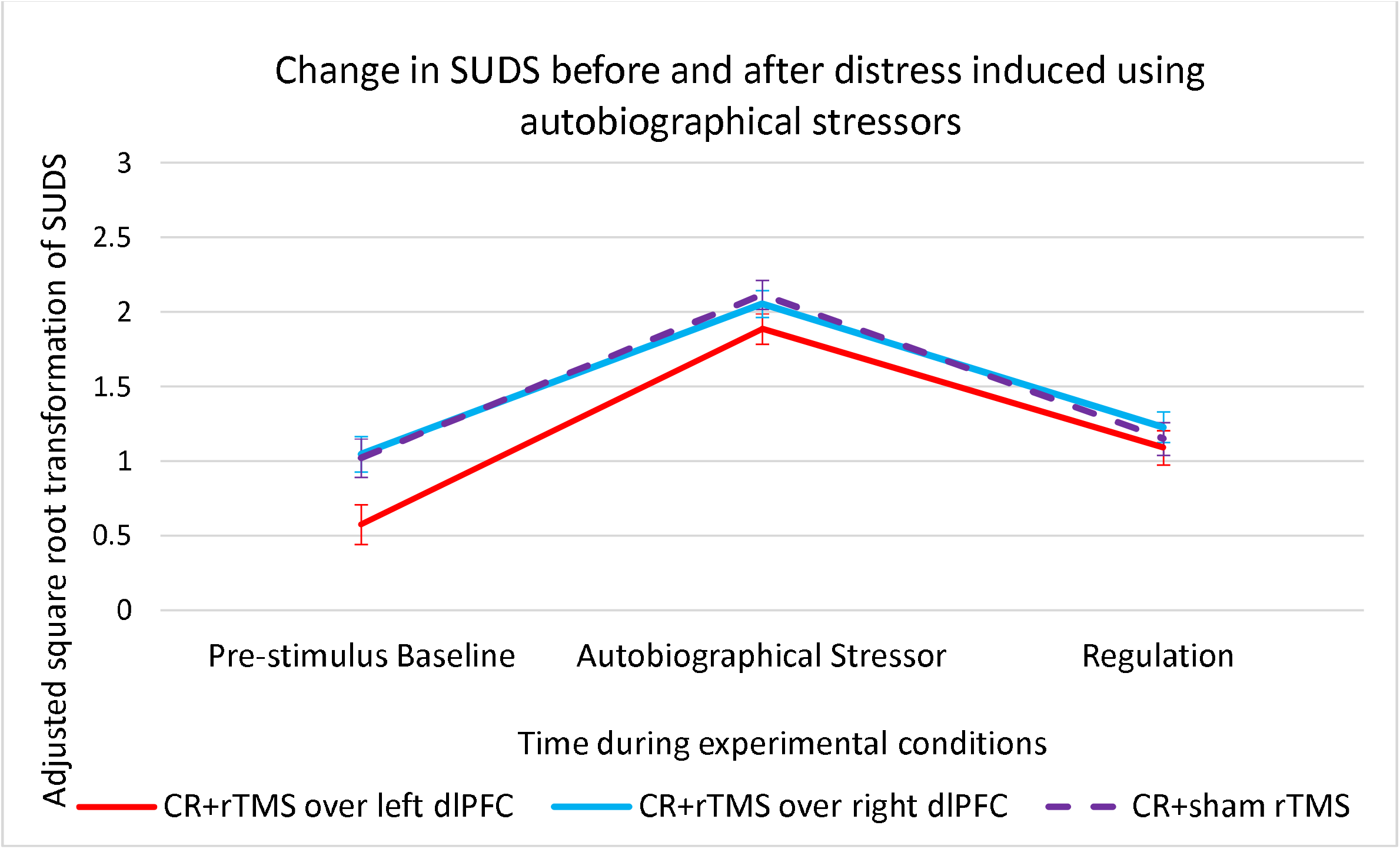
Changes in subjective units of distress (SUDS; a self-report marker of negative emotional arousal) summarized across the three experimental periods, and separated by condition, and by instruction provided. The original SUDS value was transformed using a square root function to achieve normality. Each time point on the graph represents the estimate marginal mean from the LSD pairwise comparisons from the MMANOVA main analysis which accounts for covariates. The graph presents data at the end of the pre-stimulus baseline, after the negative emotional induction using the autobiographical stressor, and at the end of the regulation period.

#### Regulation Duration Results

For the 38 participants who had above pre-stimulus baseline HR at the beginning of the regulation period, it took almost twice as long to return to HR baseline during regulation if receiving sham (M_sham_ = 129.08 s, S.D. = 194.57, N = 11) versus active neurostimulation (M_active_right_= 65.31 s, S.D. = 97.92, *n* = 15; M_active_left_ = 74.20 s, S.D. = 110.64, *n* = 12). A MMANOVA using autoregressive covariance structure (*F*_*treatment_condition*_[2, 36.48] = 3.14, *p* = .055, *p*_*FDR*_ = .10) found that that regulation duration was significantly longer in the sham condition when compared to the active right condition (*p* < .05; *d*= .62) and near significantly longer when compared to the active left condition (*p* = .08; *d* = .48). The active conditions were not significantly different (*p* = .58) and were equivalent (90% CI = [-0.495 -0.34]).

In summary, we found preliminary evidence that when compared to CR with sham neurostimulation, rTMS administered in conjunction with CR enhances physiological (HF-HRV; regulation duration) and, after left rTMS, self-reported (SUDS) indices of emotion regulation even after controlling for multiple comparisons. Left and right administration of neurostimulation leads to equivalent biological responses.

The Near-Term Effect of the Combined Intervention on Daily Distress and Use of CR Participants indicated having used CR since the previous call in 785 instances (44.73% of answered calls) and reported SUDs above 0 in 1245 calls (71.30% of answered calls). Calls happened on average every 107.06 minutes (SD = 45.08), with no difference between conditions in the amount of time that lapsed between placed calls (*p*> .05). Participants who received active left rTMS stimulation reported significantly more likelihood to use CR (EMM = 0.58, SE = 0.04) when compared to participants who received sham rTMS (EMM = 0.41, SE = 0.05) or right rTMS (EMM = 0.31, SE = 0.04), Wald χ^2^_*condition*_ (2) = 21.89, *p* < .001, *p*_FDR_ = .05.

We also found a significant main effect of treatment condition on daily distress (*F*_*treatment_condtion*_[2, 257.86] = 3.23, *p* = .04, *p*_FDR_ = .1). LSD-controlled pairwise comparisons showed that participants who received active left rTMS reported significantly less average distress during the ambulatory week (EMM_sqrt_suds_= 0.95, SE = 0.12) when compared to participants who received active right rTMS (EMM _sqrt_suds_= 1.18, SE = 0.11). The sham group (EMM_sqrt_suds_= 1.11, SE = 0.12) did not differ from the other two groups (*p*s > .05). There was no significant main effect of time or baseline (*p*s> .05).

In summary, active left rTMS led to more frequent use of CR compared to sham and right rTMS, and reduced daily distress when compared to right rTMS. The results remained significant after applying the FDR correction.

### The Long-Term Effect of the Combined Intervention

#### One Week and One Month Self-Report Results

Paired sample *t*-tests indicated that all participants significantly improved in their use of reappraisal (*t*[42] = 10.53, *d* = 1.70, *p* < .001) after the intervention. Participants were stable in their use of reappraisal from phone screen to baseline (i.e., before the intervention occurred), *t*(45) = 1.47, *p* > .05. After applying the FDR correction, there were no significant differences between treatment conditions or condition-by-time interactions for reappraisal, overall difficulties in emotion regulation, overall psychopathology severity or work and social functioning impairment (*p*s > .04; Table 2). The differences between conditions at each of the follow up time points also failed the equivalence test (90% CI included -0.5 or 0.5).

**Table 2.** Means and SDs for longitudinal outcomes for all participants and broken by condition

|  |  | **OQ-45** | **DERS** | **ERQ-R** | **WSAS** |
| --- | --- | --- | --- | --- | --- |
| **All** | Intake (n = 46) | 66.46 (17.70) | 103.89 (24.41) | 3.35 (0.81) | 16.02 (6.47) |
|  | 1 week FU (n = 43) | 58.74 (23.01) | 88.83 (25.91) | 4.67 (0.82) | 12.07 (8.58) |
|  | 1 month FU (n = 42) | 52.97 (22.56) | 82.95 (24.99) | 4.68 (0.91) | 10.95 (7.20) |
| **CR +** | Intake (n = 14) | 61.57 (19.77) | 94.57 (26.75) | 3.37 (0.94) | 15.00 (7.55) |
| **Left** | 1 week FU (n = 13) | 59.62 (24.56) | 86.92 (25.58) | 4.50 (0.99) | 14.31 (9.80) |
| **rTMS** | 1 month FU (n = 11) | 56.91 (26.30) | 84.82 (33.85) | 4.36 (1.12) | 10.27 (7.23) |
| **CR +** | Intake (n = 17) | 65.41 (18.76) | 97.88 (24.36) | 3.44 (0.81) | 16.80 (6.53) |
| **Right** | 1 week FU (n = 16) | 58.00 (26.48) | 80.68 (24.89) | 4.80 (0.89) | 10.44 (7.69) |
| **rTMS** | 1 month FU (n = 17) | 48.82 (23.86) | 76.52 (21.09) | 4.84 (0.81) | 12.29 (8.85) |
| **CR +** | Intake (n = 15) | 72.20 (13.44) | 119.40 (13.60) | 3.22 (0.73) | 16.20 (5.58) |
| **sham** | 1 week FU (n = 14) | 58.79 (18.55) | 99.93 (25.13) | 4.71 (0.58) | 11.86 (8.52) |
| **rTMS** | 1 month FU (n = 14) | 54.93 (18.24) | 89.29 (20.99) | 4.74 (0.84) | 9.86 (4.77) |
|  |  | > 63 = clinical | < 80 = | > 4.67 = | 10-20 clinical |
|  |  | impairment; | individuals not | non-clinical | impairment |
| **Normative data:** |  | reliable | in treatment |  | >20 serious |
|  |  | change = 17 |  |  | psychopatholo |
|  |  | points |  |  | gy |

*Note.* FU = follow up; CR= Cognitive restructuring training and practice on autobiographical stressors. SD = standard deviation; OQ-45 = Outcome Questionnaire-45 total score; DERS = Difficulties in Emotion Regulation Scale Total Score; ERQ-R = Emotion Regulation Questionnaire- Reappraisal Scale; WSAS = Work and Social Adjustment Scale

#### One Month Follow-Up Behavioral and Physiological Results

Participants were successful during the follow-up stressor task to increase their subjective distress from baseline (Δ_SUDSstressor-*baseline*_= 2.25, S.E. = .35, *p* < .001) and then regulate it back to baseline (Δ_SUDS_stressor-regulation_= 2.38, S.E.=.35, *p* < .001) using a fourth autobiographical stressor and CR as a regulation technique (*F*_*time*_[2, 81.55] = 26.32, *p* < .001). There were no significant differences between treatment conditions in the success of regulation when examined via either HF-HRV or SUDS (*F*_*HF-HRV*_[2, 36] *=* 1.68; *F*_*SUDS*_[2, 99.71] = 1.00; *p*s > .05). A univariate GLM test of regulation duration (M = 38.84s, SD =57.27) showed no significant difference between treatment conditions (*p* = .78; N = 29). Nevertheless, conditions were not equivalent (90% CI included -0.5 or 0.5).

In summary, participants maintained their regulation ability at follow up, but no self-reported or physiological effect of rTMS augmentation could be seen one month after its administration.

## Discussion/Conclusion

This proof-of-concept study preliminarily demonstrated the promise of augmenting emotion regulation skills training and practice with rTMS in a one-time intervention designed to target a dysregulated neural circuit. Neurostimulation over either the left or the right dlPFC enhanced immediate emotion regulation according to behavioral, physiological, and self-reported indices. The augmented gains could be seen up to one week later for those who received left dlPFC stimulation. Participants tolerated the procedures well, found the approach acceptable, and reported having a positive experience. These results are encouraging of continued efforts to combine rTMS with skills training in brief, transdiagnostic interventions.

All participants learned CR in a 45-minute session following a standardized scripted protocol and then practiced CR on their own autobiographical stressors. The session included nonspecific therapeutic factors such as positive regard, attention, and invitation to self-disclose difficulties. This training by itself proved valuable and led to a significant increase in skills use corresponding to a large effect size (Cohen’s *d* = 1.70). This result is unlikely to represent a regression to the mean but it could be attributable to the nonspecific factors present across conditions [43]. Future studies should examine the unique effects of one-time behavioral interventions on emotional dysregulation and parse out the need for skills training versus the therapeutic benefit of the participant-experimenter interaction [77].

The addition of active rTMS to CBT training enhanced emotion regulation as measured by HF-HRV, regulation duration, and self-reported distress. Active stimulation got participants to calm down faster. Sham participants achieved the same regulation effect over a longer (almost double) time frame. Therefore, neurostimulation may play an important role in enhancing the utility of the emotion regulation skill in the moment. Stimulating over the right or the left led to equivalent physiological outcomes in the moment. Beyond the intervention session, active left rTMS enhanced the acquisition of CR and reduced daily distress with noticeable effects up to a week. Given that existing protocols that combine psychotherapy and neurostimulation are 16 to 20 weeks long [30,78], our approach provides a promising novel paradigm for future research.

To test the superiority of the combination of rTMS and CR, future studies should examine whether neurostimulation alone can achieve these effects following autobiographical emotional induction without teaching and practicing an emotion regulation skill. Parsing out whether rTMS leads to reduced reactivity instead of regulation would also be an important mechanistic question to achieve additional clarity on. Furthermore, other contextual variables such as expectations given the novelty of the treatment, treatment setting, personal characteristics, placebo reactivity, and treatment history should also be examined for their relative contributions to the treatment outcomes[79].

This study fills a gap in knowledge about the utility of neuromodulation as an enhancing technique to behavioral interventions in a transdiagnostic clinical sample. We used the standard 10Hz rTMS to enhance cortical excitability in the dlPFC, in line with the findings discussed in the introduction that point to transdiagnostic hypoactivity in this region during emotion regulation tasks. We also employed neuronavigation using the 10-20 system. This approach provides a more precise localization of the dlPFC when compared to the more traditional 5 cm rule [80]. We opted to divide the rTMS session into 4 epochs in order to make engagement in autobiographical emotion regulation feasible. This decision was aligned with prior research highlighting that breaks may enhance facilitatory after-effects [81]. Furthermore, having participants spend more than 10 minutes in effortful emotion regulation would be difficult to accomplish and receiving rTMS during emotional induction would dampen the emotional experience [82], reducing the opportunity for regulation. Our results demonstrate that these decisions did not alter the potential of neurostimulation to enhance behavioral training and that therapeutic activities, like skills practice, could take place during rTMS successfully.

The study had some important limitations including the sample size and lack of matched control group for the skills training component. The study was powered expecting a moderate to large effect size and we found small to moderate effect sizes, which may suggest that the results are underpowered. A more stringent approach to defining low CR would have been to only include those who score 2 SD below the pooled mean. Targeting for rTMS in this study used exterior head measurements and, therefore, may have not resulted in excitation of a relevant node of the emotion regulation neural network. The interval between training sessions was short, which may reduce the effectiveness of skills training [83]. The combination of rTMS with a spaced training or irregular training protocol may optimize and enhance learning [83]. Finally, It is important to highlight that bias may exist in the publication of positive neurostimulation results [84] and that additional scrutiny may be needed for this experimental approach.

The intervention design could also benefit from hypothesis driven optimization. Several parameters that can improve the efficacy of rTMS have been identified and should be tested in future iterations of this intervention: targeting networks versus regions[85-87], targeting using functional MRI with neuro-navigation[20,88], adjusting the inter-trial interval[88,89], and using E-field modeling [90]. In addition, maximizing the context dependent nature of rTMS by identifying the optimal time for emotion induction and for neurostimulation onset could also enhance efficacy. On the behavioral side, refinement of learning parameters such as the spacing between regulation trials to optimize synaptic plasticity[83], accounting for nonspecific factors, and ensuring successful skills acquisition and implementation could further enhance the efficacy of the combined intervention.

In summary, our goal was to begin the development of a time-limited combined intervention for transdiagnostic emotional dysregulation. We found preliminary evidence that rTMS augments behavioral skills training in the moment, and that the one-time session of combined rTMS-CR intervention may lead to changes in synaptic plasticity and have effects that last as long as a week. Our findings provide a promising first step in the development of novel neuroscience-driven treatment paradigms that address this hard-to-treat, transdiagnostic clinical population.

## Data Availability

No external datasets or supplemental materials are available at this time. Data sets and protocol materials can be obtained by e-mailing the first author

## Statements

## Acknowledgements (optional)

Data in the present paper were also presented as part of several conference talks. The authors would like to thank the participants and to acknowledge Lisalynn Kelley, Zach Rosenthal, PhD, Caitlin Fang, PhD, Kevin Haworth, PhD, Megan Renna, PhD, Paul Geiger, PhD, Simon Davis, PhD, Moria Smoski, PhD, Tim Strauman, PhD, Michael Babyak, PhD our research assistants and DUHS staff for their contributions.

## Statement of Ethics

### Study approval statement

The study was approved by the Duke University Health System Institutional Review Board (approval number: Pro00066383) and pre-registered on ClinicalTrials.gov (NCT02573246).

### Consent to participate statement

Written informed consent was obtained before any study procedures began.

## Author Contributions

All authors contributed significantly to the present manuscript. Dr. Neacsiu conceptualized and conducted the study. Drs. LaBar, Appelbaum, and Lisanby provided design input, helped problem solve study related issues throughout the study, and contributed to the manuscript preparation. Dr. Beynel conducted the neurostimulation procedures. Dr. Powers completed our physiological preprocessing pipeline. Dr. Szabo served as the study doctor throughout the study.

## Data Availability Statement

All data is available upon request.

## References

1. Gross JJ. Handbook of Emotion Regulation, 2nd ed. New York: The Guilford Press; 2013, 669 p.

2. Kring AM, Werner KH. Emotion Regulation and Psychopathology. In: Philippot P, Feldman RS, editors. The regulation of emotion. Mahwah, NJ: Lawrence Erlbaum Associates Publishers; 2004. p. 359–85.

3. Wolff JC, Thompson E, Thomas SA, Nesi J, Bettis AH, Ransford B, et al. Emotion dysregulation and non-suicidal self-injury: A systematic review and meta-analysis. Eur Psychiatry 2019;59:25–36.

4. Bekh BD, DeFife JA, Guarnaccia C, Phifer MJ, Fani MN, Ressler KJ, et al. Emotion dysregulation and negative affect: association with psychiatric symptoms. J Clin Psychiatry 2011;72(5):685–91.

5. Fernandez KC, Jazaieri H, Gross JJ. Emotion Regulation: A Transdiagnostic Perspective on a New RDoC Domain. Cognit Ther Res 2016;40(3):426–40.

6. Campbell-Sills L, Barlow DH. Incorporating Emotion Regulation into Conceptualizations and Treatments of Anxiety and Mood Disorders. In: Gross JJ, editor. Handbook of emotion regulation. New York, NY, US: Guilford Press; 2007. p. 542–59.

7. Gaher RM, Hofman NL, Simons JS, Hunsaker R. Emotion Regulation Deficits as Mediators Between Trauma Exposure and Borderline Symptoms. Cogn Ther Res 2013;37(3):466–75.

8. Gratz KL, Levy R, Tull MT. Emotion Regulation as a Mechanism of Change in an Acceptance-Based Emotion Regulation Group Therapy for Deliberate Self-Harm Among Women With Borderline Personality Pathology. J Cogn Psychotherapy 2012;26(4):365–80.

9. Barlow DH, Farchione TJ, Bullis JR, Gallagher MW, Murray-Latin H, Sauer-Zavala S, et al. The Unified Protocol for Transdiagnostic Treatment of Emotional Disorders Compared With Diagnosis-Specific Protocols for Anxiety Disorders A Randomized Clinical Trial. JAMA Psychiatry 2017;74(9):875–84.

10. Neacsiu AD, Eberle JW, Kramer R, Wiesmann T, Linehan MM. Dialectical behavior therapy skills for transdiagnostic emotion dysregulation: A pilot randomized controlled trial. Behav Res Ther 2014;59:40–51.

11. Berking M, Ebert D, Cuijpers P, Hofmann SG. Emotion Regulation Skills Training Enhances the Efficacy of Inpatient Cognitive Behavioral Therapy for Major Depressive Disorder: A Randomized Controlled Trial. Psychotherapy and Psychosomatics 2013;82(4):234–45.

12. Farchione TJ, Fairholme CP, Ellard KK, Boisseau CL, Thompson-Hollands J, Carl JR, et al. Unified protocol for transdiagnostic treatment of emotional disorders: a randomized controlled trial. Behav Ther 2012;43(3):666–78.

13. Linehan MM, Korslund KE, Harned MS, Gallop RJ, Lungu A, Neacsiu AD, et al. Dialectical behavior therapy for high suicide risk in individuals with borderline personality disorder: a randomized clinical trial and component analysis. JAMA Psychiatry 2015;72(5):475–82.

14. Neacsiu AD, Bohus M, Linehan MM. Dialectical Behavior Therapy: An Intervention for Emotion Dysregulation. In: Gross, JJ, editor. Handbook of emotion regulation. 2nd edition. New York, NY: The Guilford Press; 2013. p. 491–508.

15. Cuijpers P, Cristea IA, Karyotaki E, Reijnders M, Hollon SD. Component studies of psychological treatments of adult depression: a systematic review and meta-analysis. Psychother Res 2019;29(1):15–29.

16. Roth A, Fonagy P. What works for whom? A critical review of psychotherapy research. New York, NY: The Guilford Press; 2005.

17. Kazdin AE, Blase SL. Rebooting Psychotherapy Research and Practice to Reduce the Burden of Mental Illness. Perspect Psychol Sci 2011;6(1):21–37.

18. O’Neill A, Frodl T. Brain structure and function in borderline personality disorder. Brain Struct Funct 2012;217(4):767–82.

19. Zilverstand A, Parvaz MA, Goldstein RZ. Neuroimaging cognitive reappraisal in clinical populations to define neural targets for enhancing emotion regulation. A systematic review. NeuroImage. 2017;151:105–16.

20. Neacsiu AD, Lisanby SH. Magnetic stimulation for depression: Subconvulsive and convulsive approaches. Neuromodulation in Psychiatry. 2015:155–80.

21. Beynel L, Powers JP, Appelbaum LG. Effects of repetitive transcranial magnetic stimulation on resting-state connectivity: A systematic review. NeuroImage. 2020;211(116596) 1:11.

22. De Raedt R, Vanderhasselt M-A, Baeken C. Neurostimulation as an intervention for treatment resistant depression: From research on mechanisms towards targeted neurocognitive strategies. Clin Psychol Rev 2015;41:61–9.

23. De Raedt R, Leyman L, Baeken C, Van Schuerbeek P, Luypaert R, Vanderhasselt MA, et al. Neurocognitive effects of HF-rTMS over the dorsolateral prefrontal cortex on the attentional processing of emotional information in healthy women: An event-related fMRI study. Biol Psychology 2010;85(3):487–95.

24. Feeser M, Prehn K, Kazzer P, Mungee A, Bajbouj M. Transcranial Direct Current Stimulation Enhances Cognitive Control During Emotion Regulation. Brain Stimulation 2014;7(1):105–12.

25. Luber B, Steffener J, Tucker A, Habeck C, Peterchev AV, Deng ZD, et al. Extended remediation of sleep deprived-induced working memory deficits using fMRI-guided transcranial magnetic stimulation. Sleep. 2013;36(6):857–71.

26. Van den Eynde F, Claudino AM, Mogg A, Horrell L, Stahl D, Ribeiro W, et al. Repetitive transcranial magnetic stimulation reduces cue-induced food craving in bulimic disorders. Biol Psychiatry 2010;67(8):793–5.

27. Trevizol AP, Shiozawa P, Cook IA, Sato IA, Kaku CB, Guimarães FB, et al. Transcranial magnetic stimulation for obsessive-compulsive disorder: an updated systematic review and meta-analysis. J ECT 2016;32(4):262–6.

28. Gorelick DA, Zangen A, George MS. Transcranial magnetic stimulation (TMS) in the treatment of substance addiction. Ann N Y Acad Sci 2014;1327(1):79–93.

29. Neacsiu AD, Luber BM, Davis SW, Bernhardt E, Strauman TJ, Lisanby SH. On the Concurrent Use of Self-System Therapy and Functional Magnetic Resonance Imaging-Guided Transcranial Magnetic Stimulation as Treatment for Depression. J ECT 2018;34(4):266–73.

30. Kozel FA, Motes MA, Didehbani N, DeLaRosa B, Bass C, Schraufnagel CD, et al. Repetitive TMS to augment cognitive processing therapy in combat veterans of recent conflicts with PTSD: a randomized clinical trial. J Affect Disord 2018;229:506–14.

31. Cunningham DA, Varnerin N, Machado A, Bonnett C, Janini D, Roelle S, et al. Stimulation targeting higher motor areas in stroke rehabilitation: A proof-of-concept, randomized, double-blinded placebo-controlled study of effectiveness and underlying mechanisms. Restor Neurol Neurosci 2015;33(6):911–26.

32. Ochsner KN, Bunge SA, Gross JJ, Gabrieli JDE. Rethinking feelings: an FMRI study of the cognitive regulation of emotion. J Cogn Neurosci 2002;14(8):1215–29.

33. Webb TL, Miles E, Sheeran P. Dealing with feeling: a meta-analysis of the effectiveness of strategies derived from the process model of emotion regulation. Psychol Bull 2012;138(4):775–808.

34. Gross JJ, Jazaieri H. Emotion, emotion regulation, and psychopathology: An affective science perspective. Clin Psychol Sci 2014;2(4):387–401.

35. Ochsner KN, Silvers JA, Buhle JT. Functional imaging studies of emotion regulation: a synthetic review and evolving model of the cognitive control of emotion. Ann N Y Acad Sci 2012;1251:E1–24.

36. Powers JP, LaBar KS. Regulating emotion through distancing: A taxonomy, neurocognitive model, and supporting meta-analysis. Neurosci Biobehav Rev 2019;96:155–73.

37. Thayer JF, Åhs F, Fredrikson M, Sollers III JJ, Wager TD. A meta-analysis of heart rate variability and neuroimaging studies: implications for heart rate variability as a marker of stress and health. Neurosci Biobehav Rev 2012;36(2):747–56.

38. Lane RD, McRae K, Reiman EM, Chen K, Ahern GL, Thayer JF. Neural correlates of heart rate variability during emotion. NeuroImage. 2009;44(1):213–22.

39. Butler EA, Wilhelm FH, Gross JJ. Respiratory sinus arrhythmia, emotion, and emotion regulation during social interaction. Psychophysiology. 2006;43(6):612–22.

40. Demaree HA, Robinson JL, Everhart DE, Schmeichel BJ. Resting RSA is associated with natural and self-regulated responses to negative emotional stimuli. Brain Cogn 2004;56(1):14–23.

41. Elliot AJ, Payen V, Brisswalter J, Cury F, Thayer JF. A subtle threat cue, heart rate variability, and cognitive performance. Psychophysiology. 2011;48(10):1340–5.

42. Fenton-O’Creevy M, Lins JT, Vohra S, Richards DW, Davies G, Schaaff K. Emotion regulation and trader expertise: Heart rate variability on the trading floor. J Neurosci Psychol Econ 2012;5(4):227–37.

43. Guidi J, Brakemeier E-L, Bockting CL, Cosci F, Cuijpers P, Jarrett RB, et al. Methodological recommendations for trials of psychological interventions. Psychother Psychosom 2018;87(5):276–84.

44. Faul F, Erdfelder E, Buchner A, Lang AG. Statistical power analyses using G*Power 3.1: tests for correlation and regression analyses. Behav Res Methods 2009;41(4):1149–60.

45. Geisler FC, Kubiak T. Heart rate variability predicts self-control in goal pursuit. Eur J Per 2009;23(8):623–33.

46. First MB, Williams JB, Karg RL. Structured Clinical Interview for DSM-5 Disorders (Research Version: SCID-5-RV). Washington, DC: American Psychiatric Publishing; 2015.

47. First MB, Williams JB, Benjamin LS, Spitzer MD. Structured Clinical Interview for DSM-5 Personality Disorders (SCID-5-PD). Washington D.C.: American Psychiatric Association Publishing; 2016.

48. Pitman RK, Orr SP, Forgue DF, de Jong JB, Claiborn JM. PSychophysiologic assessment of posttraumatic stress disorder imagery in vietnam combat veterans. Arch Gen Psychiatry 1987;44(11):970–5.

49. Dunn LM. PPVT-revised manual. Circle Pines, MN: American Guidance Service; 1981.

50. Scott NW, McPherson GC, Ramsay CR, Campbell MK. The method of minimization for allocation to clinical trials. a review. Control Clin Trials 2002;23(6):662–74.

51. Taves DR. Minimization: a new method of assigning patients to treatment and control groups. Clin Pharmacol Ther 1974;15(5):443–53.

52. Linehan M. DBT Skills Training Manual.2nd ed. New York, NY:Guilford Publications; 2014.

53. Beck AT, Emery G, Greenberg RL. Anxiety disorders and phobias: A cognitive perspective. Basic Books; 2005.

54. Shurick AA, Hamilton JR, Harris LT, Roy AK, Gross JJ, Phelps EA. Durable effects of cognitive restructuring on conditioned fear. Emotion. 2012;12(6):1393–7.

55. Stiglmayr C, Schmahl C, Bremner JD, Bohus M, Ebner-Priemer U. Development and psychometric characteristics of the DSS-4 as a short instrument to assess dissociative experience during neuropsychological experiments. Psychopathology. 2009;42(6):370–4.

56. Beam W, Borckardt JJ, Reeves ST, George MS. An efficient and accurate new method for locating the F3 position for prefrontal TMS applications. Brain Stimulation 2009;2(1):50–4.

57. Rossi S, Hallett M, Rossini PM, Pascual-Leone A, Group SoTC. Safety, ethical considerations, and application guidelines for the use of transcranial magnetic stimulation in clinical practice and research. Clin Neurophysiol 2009;120(12):2008–39.

58. Rossi S, Antal A, Bestmann S, Bikson M, Brewer C, Brockmöller J, et al. Safety and recommendations for TMS use in healthy subjects and patient populations, with updates on training, ethical and regulatory issues: Expert Guidelines. Clin Neurophysiol 2021; 132 (1): 269–306.

59. Wassermann EM. Risk and safety of repetitive transcranial magnetic stimulation: report and suggested guidelines from the International Workshop on the Safety of Repetitive Transcranial Magnetic Stimulation, June 5–7, 1996. Electroencephalogr Clin Neurophysiol 1998;108(1):1–16.

60. Chou YH, You H, Wang H, Zhao YP, Hou B, Chen NK, et al. Effect of repetitive transcranial magnetic stimulation on fMRI resting-state connectivity in multiple system atrophy. Brain connectivity 2015;5(7):451–9.

61. Berntson GG, Thomas Bigger Jr J, Eckberg DL, Grossman P, Kaufmann PG, Malik M, et al. Heart rate variability: origins, methods, and interpretive caveats. Psychophysiology. 1997;34(6):623–48.

62. Gratz KL, Roemer L. Multidimensional Assessment of Emotion Regulation and Dysregulation: Development, Factor Structure, and Initial Validation of the Difficulties in Emotion Regulation Scale. J Psychopathol Behav Assess 2004;26(1):41–54.

63. Lambert MJ, Burlingame GM, Umphress V, Hansen NB, Vermeersch DA, Clouse GC, et al. The reliability and validity of the Outcome Questionnaire. Clin Psychol Psychother: Intl J Theory Practice. 1996;3(4):249–58.

64. Mundt JC, Marks IM, Shear MK, Greist JM. The Work and Social Adjustment Scale: a simple measure of impairment in functioning. Br J Psychiatry 2002;180(5):461–4.

65. Verbeke G. Linear mixed models for longitudinal data. Linear mixed models in practice: Springer; 1997. p. 63–153.

66. Feingold A. Effect sizes for growth-modeling analysis for controlled clinical trials in the same metric as for classical analysis. Psychol Methods 2009;14(1):43–53.

67. Cohen J. Statistical power analysis for the behavioral sciences. Revised. New York, NY: Academic Press; 1977.

68. Benjamini Y, Hochberg Y. Controlling the false discovery rate: a practical and powerful approach to multiple testing. J Royal statistical society: series B (Methodological). 1995;57(1):289–300.

69. Liang K-Y, Zeger SL. Longitudinal data analysis using generalized linear models. Biometrika. 1986;73(1):13–22.

70. Raudenbush SW, Bryk AS. Hierarchical linear models: Applications and data analysis methods. London, UK: Sage publications; 2002.

71. Zeger SL, Liang K-Y, Albert PS. Models for longitudinal data: a generalized estimating equation approach. Biometrics. 1988:1049–60.

72. Zeger SL, Liang K-Y. Longitudinal data analysis for discrete and continuous outcomes. Biometrics. 1986:121–30.

73. Walker E, Nowacki AS. Understanding equivalence and noninferiority testing. J Gen Intern Med 2011;26(2):192–6.

74. Lakens D, Scheel AM, Isager PM. Equivalence testing for psychological research: A tutorial. Adv Methods Practices Psychol Sci 2018;1(2):259–69.

75. Leichsenring F, Luyten P, Hilsenroth MJ, Abbass A, Barber JP, Keefe JR, et al. Psychodynamic therapy meets evidence-based medicine: a systematic review using updated criteria. Lancet Psychiatry 2015;2(7):648–60.

76. Norman GR, Sloan JA, Wyrwich KW. Interpretation of changes in health-related quality of life: the remarkable universality of half a standard deviation. Med Care 2003; 41(5):582–92.

77. Wodack KH, Poppe AM, Tomkotter L, Bachmann KA, Strobel CM, Bonk S, et al. Individualized early goal-directed therapy in systemic inflammation: is full utilization of preload reserve the optimal strategy? Crit Care Med. 2014;42(12):e741–51.

78. Kozel FA, Van Trees K, Larson V, Phillips S, Hashimie J, Gadbois B, et al. One hertz versus ten hertz repetitive TMS treatment of PTSD: a randomized clinical trial. Psych Res 2019;273:153–62.

79. Fava GA, Guidi J, Rafanelli C, Rickels K. The clinical inadequacy of the placebo model and the development of an alternative conceptual framework. Psychother Psychosom 2017;86(6):332–40.

80. McClintock SM, Reti IM, Carpenter LL, McDonald WM, Dubin M, Taylor SF, et al. Consensus recommendations for the clinical application of repetitive transcranial magnetic stimulation (rTMS) in the treatment of depression. J Clin Psych 2017;79(1):16cs10905:35-48.

81. Rothkegel H, Sommer M, Paulus W. Breaks during 5 Hz rTMS are essential for facilitatory after effects. Clin Neurophysiol 2010;121(3):426–30.

82. Remue J, Vanderhasselt M-A, Baeken C, Rossi V, Tullo J, De Raedt R. The effect of a single HF-rTMS session over the left DLPFC on the physiological stress response as measured by heart rate variability. Neuropsychology. 2016;30(6):756–66.

83. Smolen P, Zhang Y, Byrne JH. The right time to learn: mechanisms and optimization of spaced learning. Nat Rev Neurosci 2016;17(2):77–88.

84. Amad A, Jardri R, Rousseau C, Larochelle Y, Ioannidis JP, Naudet F. Excess significance bias in repetitive transcranial magnetic stimulation literature for neuropsychiatric disorders. Psychother Psychosom 2019;88(6):363–70.

85. Opitz A, Fox MD, Craddock RC, Colcombe S, Milham MP. An integrated framework for targeting functional networks via transcranial magnetic stimulation. NeuroImage. 2016;127:86–96.

86. Fox MD, Buckner RL, White MP, Greicius MD, Pascual-Leone A. Efficacy of transcranial magnetic stimulation targets for depression is related to intrinsic functional connectivity with the subgenual cingulate. Biol Psychiatry 2012;72(7):595–603.

87. Fox MD. Mapping symptoms to brain networks with the human connectome. N Engl J Med 2018;379(23):2237–45.

88. Beynel L, Appelbaum LG, Luber B, Crowell CA, Hilbig SA, Lim W, et al. Effects of online repetitive transcranial magnetic stimulation (rTMS) on cognitive processing: A meta-analysis and recommendations for future studies. Neurosci Biobehav Rev 2019;107:47–58.

89. Cash RF, Dar A, Hui J, De Ruiter L, Baarbé J, Fettes P, et al. Influence of inter-train interval on the plastic effects of rTMS. Brain Stimulation 2017;10(3):630–6.

90. Deng ZD, Lisanby SH, Peterchev AV. Electric field depth–focality tradeoff in transcranial magnetic stimulation: simulation comparison of 50 coil designs. Brain Stimulation 2013;6(1):1–13.

