## Supplementary Materials for "Enhancing Cognitive Restructuring with Concurrent Repetitive Transcranial Magnetic Stimulation: A Transdiagnostic Randomized Controlled Trial"

***Supplemental Materials***

#### Supplemental Background Information

Because of its contribution to disorders such as affective, stress, and personality disorders, emotional dysregulation is a serious public health concern that causes an enormous burden on society and its economy[1-4]. The intensive burden on patients to come into an office for multiple sessions and put significant effort into learning new skills leads to treatment dropout and avoidance of psychotherapies, especially for emotionally dysregulated adults[5-7].

Experiments probing the utility of neurostimulation on emotional dysregulation have only been conducted with non-clinical samples and have not yet been translated to interventions. An exception is a study where 10 sessions of 10-Hz rTMS over the right dlPFC led to improvements in affective instability and anger dysregulation in a borderline personality disorder (BPD) sample[8]. Nevertheless, this study did not include a comparison with a control treatment and did not have a longitudinal assessment of effects, leaving gaps in what could be inferred from the results. Therefore, the effectiveness of rTMS on clinical emotional dysregulation is unknown. Furthermore, rTMS as a stand-alone treatment is unlikely to solve the burden barrier that psychotherapies have. Successful stand-alone rTMS treatments require 10-20 daily sessions in a medical office, which can be equally burdensome[9]. Studies that use rTMS as a potentiation tool have shown beneficial effects of both left and right dlPFC stimulation[10-14].

In addition to subjective reports of affect, we collected high frequency heart rate variability (HF-HRV) as an objective physiological measure of emotion regulation that could be examined in the moment without interfering with skill practice and neurostimulation. In the context of emotional arousal, HF-HRV is positively correlated with dlPFC or mPFC activation (z-scores > 2.95)[15,16] and is a sensitive measure of adaptive emotion regulation[17-19]. Participants who engage in effective emotion regulation have higher changes in HF-HRV from baseline than those who do not engage in regulation[20,21]. HF-HRV has been especially connected with effective and not with maladaptive regulation[22,23]. Therefore, we computed HF-HRV and interpreted increases in this measure as indicative of an increase in effective regulation.

#### Supplemental Methods and Materials

**Participants**

Participants were recruited between 4/27/2016 and 1/3/2019 through online websites (e.g., Craigslist), flyers, and physician referrals. We aimed to enroll 20 participants in each condition to align with other neurostimulation studies where 6-20 adults per condition were sufficient to demonstrate proof-of-concept for novel treatments [24,25] and following power analyses. Interested participants completed an online screen and were subsequently called for additional screening if they did not report meeting any exclusion criteria. Participants were recruited in tandem for two studies: the main study presented in this paper (N = 83 participants enrolled), as well as a supplementary study that examined neural effects of our intervention (N = 65 participants enrolled). Participants were assigned to one study or the other depending on their qualification and interest in the two studies, and if they qualified for both they were randomized (coin toss) to one or the other.

Of 63 treatment-seeking community participants who were screened and eligible, 47 participants were randomized. The first randomized participant piloted all procedures and their data was discarded. The rest of 46 participants are considered intent-to-treat (ITT). Participants were transdiagnostic and met criteria for at least one of the following diagnoses according to the Structured Clinical Interview (SCID-5) for *DSM-5* Disorders [26]: major depressive disorder (MDD; *n* = 9), persistent depressive disorder (PDD; *n* = 5), premenstrual dysphoric disorder (PMDD; *n =* 6), bipolar II disorder (*n* = 1), other depressive disorder (*n* = 2), social anxiety disorder (SAD; *n* = 17), generalized anxiety disorder (GAD; *n* = 18), specific phobia (SP; *n =* 10), other specified anxiety disorder (OASD; *n* = 6), obsessive compulsive disorder (OCD; *n* = 2), body dysmorphic disorder (BDD; *n =* 3), hoarding disorder (*n* = 1), trichotillomania (*n* = 1), excoriation disorder (*n* = 4), bulimia nervosa (BN; *n* = 1), other eating disorder (*n* = 2), somatic symptom disorder (*n* = 1), illness anxiety disorder (*n* = 1), adult attention deficit hyperactivity disorder (ADHD; *n* =7), intermittent explosive disorder (IEE; *n* = 3), acute stress disorder (*n* = 2), posttraumatic stress disorder (PTSD; *n* = 1), other stress disorder (*n* = 6), adjustment disorder (*n* = 5). Participants met criteria for an average of 2.53 (*SD =* 1.65) current diagnoses and 4.20 (*SD* = 1.94) lifetime diagnoses, and 15 participants (31.92%) met criteria for at least one personality disorder according to the SCID-5-PD [27]. Of note, a transdiagnostic sample is defined using a different set of criteria than DSM diagnostics [28], an approach is highly encouraged by the NIMH RDoC framework [29].

Low use of CR was operationally defined as a mean score lower than 4.7 on the cognitive reappraisal subscale (range: 1-7) of the Emotion Regulation Questionnaire (ERQ)[30] at the intake assessment. This cutoff was computed by pooling ERQ reappraisal means from 18 studies (N = 4331 participants) published before 2014 that examined the ERQ on US samples [30-47]. The pooled mean across these studies was 4.70 (*SD_pooled_* = 0.99). An average score of 4.7 or lower indicates that participants were mostly neutral or disagreed with statements about using cognitive restructuring to change their emotions. We considered adults who scored below this pooled mean to have “low use” of CR strategies and therefore to be the optimal candidates for a one-time CR intervention. ITT participants scored an average of 3.35 (SD = 0.81) on the ERQ cognitive reappraisal subscale, with no significant difference in scores between treatment conditions (*F*(2, 43) = 0.29, *p* = .75).

Participants were excluded if they were younger than 18 and older than 65; scored above the ERQ reappraisal cutoff; did not report any mental health difficulties; endorsed current substance or alcohol abuse or a history of psychosis; were currently in (or planning to start) CBT; needed immediate hospitalization; were at high risk for suicide; or were at increased risk for seizure during neuromodulation because of medication, neurological disorder, brain injury, or family history. Pregnant women, homeless individuals, individuals who could not travel to Duke, who did not understand English, or who had low verbal IQ [48] and could not understand the intervention were also excluded. Participants unwilling to answer the ambulatory calls were also excluded. (See Figure 1 for additional details.) No changes to inclusion/exclusion were done after beginning enrollment.

Participants received $150 compensation for all study procedures. Additional funds were awarded after the study start for a supplement which was reported under the same clinical trial ID but was conducted as an independent study. The data from the supplement are not included in this paper. The study enrollment and follow up was closed on 1/14/2019, at the conclusion of the funding period.

#### Procedures

This study involved a total of three sessions: an intake session, the intervention session combining rTMS with skills training, and a follow-up session one month after intervention. Participants also completed an ambulatory assessment of distress and CR use for a week following the intervention (8x per day for 7 days), and a battery of self-reports remotely via an online link at the end of that week. The following sections describe each of these sessions.

##### Intake Session

After providing voluntary, written informed consent, participants completed diagnostic assessments (SCID-5) and a questionnaire packet. Qualified participants underwent an interview to identify four autobiographical stressors to be used for emotional induction. Using the method developed by Pitman and colleagues [49] and used in studies of adults with affective disorders [50], we created four negative emotional arousal scripts, three of which were used in a random order during the intervention day and one during the 1-month follow-up assessment. In brief, the assessor asked the participant to describe three moderate stressors from the past month, as well as a stressor that tended to reoccur for them. Participants wrote a description of each event, and the assessor worked with the participant to establish a clear story for each event that could be recited in 30-40 seconds. Following the assessment, these scripts were narrated by the PI and digitally recorded into .wav audio files to be presented during the subsequent testing sessions. The PI decided whom to enroll in the study and was kept blind to treatment condition assignment until the end of the study. All procedures were conducted at DUHS core facilities.

###### **Randomization**

Randomization was done using a minimization algorithm [51,52] using the QMinim software (<http://rct.mui.ac.ir/q/index.php>) by an independent staff member (Ms. Kelley) and kept blind from the assessor, PI and participant. A parallel clinical trial design was used with equal ratio assignment to each condition. The randomization information was saved in a password protected database to which only Ms. Kelley had the password. Two sealed envelopes were created for each participant, one to be opened at the end of the study by the assessor and participant to discuss randomization, and one to be opened by the TMS technician who set up the coil before the intervention for either the sham or active paradigm. Sham participants were further randomized to receive TMS to either the right or left DLPFC. From the 15 participants in the sham treatment condition, 7 had the TMS coil placed over the left dlPFC and 8 over the right dlPFC.

##### Intervention Session

Participants returned for the 3.5-hour intervention session within a month of intake.

**Behavioral Intervention.** The first 45-60 min of this session was spent on skills training, one-on-one with the first author or a trained psychologist, and was focused on learning CR and practicing on standardized and personal examples. Skills training used standardized procedures blending psychotherapeutic approaches [53,54] with instructions in CR that matched prior neuroimaging studies [55]. Participants were told that one validated way to change emotional experiences is by thinking differently about the situation that prompts the emotion [54,56-58] or by reorganizing the cognitive elements involved in the emotion [59].

Following Gross’s model [60], we defined CR as interpreting emotional stimuli in a way in which the target emotional relevance is de-emphasized or the interpretation leads to a different emotion [61]. First, participants were taught to adopt a detached and unemotional attitude as they thought about their autobiographical situation [62]. Detachment was instructed by examining the situation with objectivity or putting spatial or temporal distance between the current moment and the situation. We called this strategy **distancing** [63].

Second, participants were taught **reframing** using an adapted version of existing paradigms [55,64,65]. Specifically, we emphasized with pictures and examples the relationship between thoughts and emotions, identified helpful ways of thinking, and instructed participants to find interpretations that are less toxic in order to be effective rather than right when upset. Participants learned to ask about elements of the situation that they did not pay attention to or information that was missing, and to reframe their cognitions based on the full picture with an eye towards reducing distress. Participants were also taught to examine the worst-case scenario, the probability of it occurring, and the likelihood of survival if it occurred.

Functional neuroimaging studies examining CR have found activation distributed throughout the frontolimbic emotion regulation network, including the dlPFC, vlPFC, ACC, mPFC/frontal pole, amygdala, and insula [63,66,67]. Of these regions, the dlPFC is a critical node in the effective use of cognitive emotion regulation [68] that is also the most accessible site for neuromodulation with good demonstration of patient tolerance [9]. We define effective regulation as a process that reduces unwanted arousal without an unintended consequence or cost [69,70]. In other words, effectiveness of regulation entails successfully reducing arousal back to baseline, and doing so by using a strategy that is deemed effective (e.g., restructuring, problem solving, breathing). An ineffective strategy is costly or brings negative consequences (drinking, rumination, suppression). CR is expected to be an effective strategy.

The CR training ended with a quiz that included definitions as well as hypothetical scenarios for practice purposes. It is important to highlight that our behavioral intervention was not a therapy session; rather we designed it to be a skills training session. Given that psychotherapy sessions may vary in their unique success [71,72], we wanted to have a metric of success for this session before proceeding with the rest of the intervention. An experienced psychotherapist conducted the skills training, where a skill the participants did not know well was taught, and practice on generalized and personalized examples was engaged in. We measured “success” by asking participants to pass a quiz that demonstrated at least some knowledge of the terms used. As we went through examples with participants, we added as many examples as needed to ensure that participants “got the skill”.

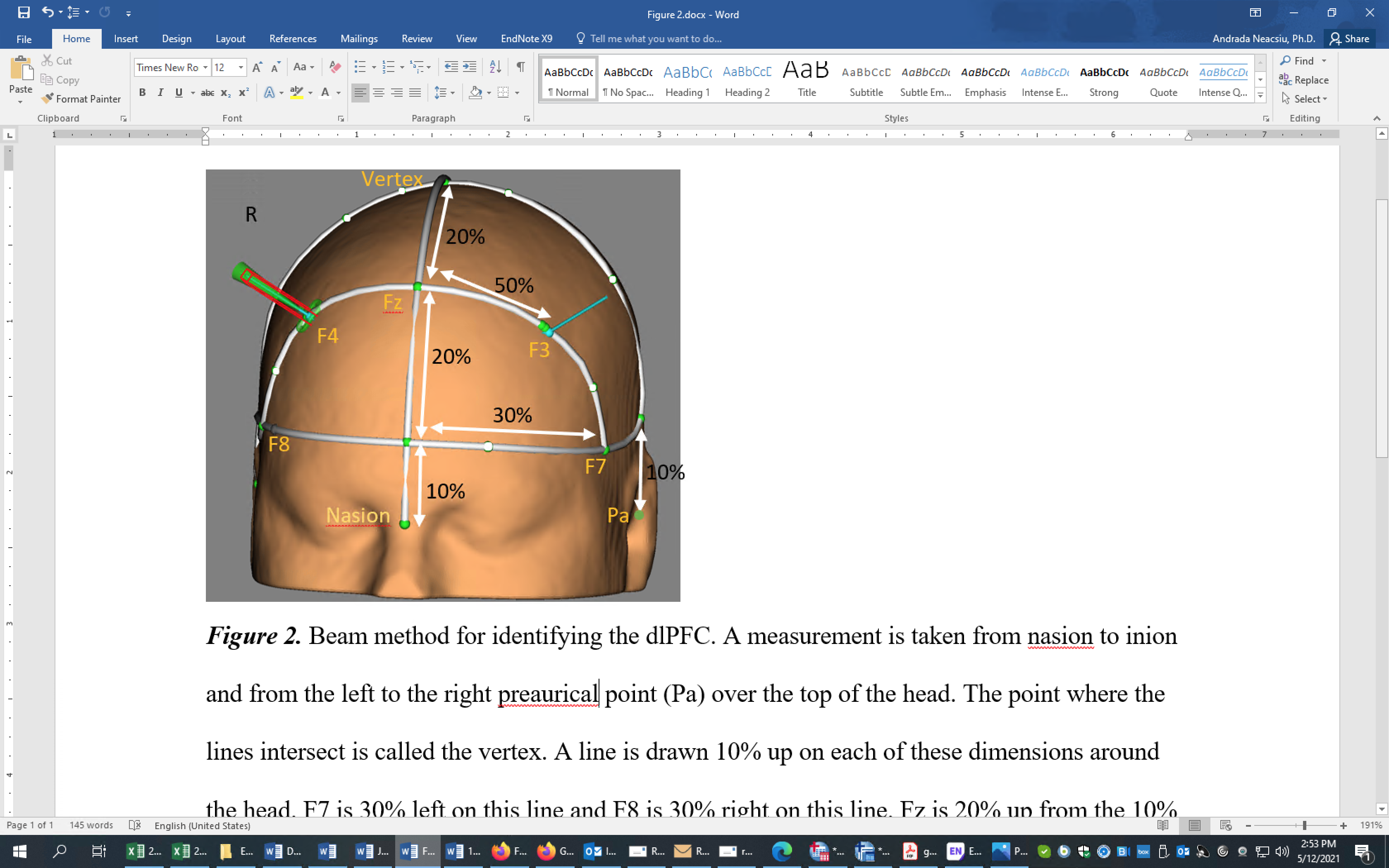

*Supplementary Figure 1.* Beam method for identifying the dlPFC. A measurement is taken from nasion to inion (not shown) and from the left to the right preaurical point (Pa) over the top of the head. The point where the lines intersect is called the vertex. A line is drawn 10% up on each of these dimensions around the head. F7 is 30% left on this line and F8 is 30% right on this line. Fz is 20% up from the 10% line on the nasion-inion line. F3 is at the half of the distance between Fz and F7. F4 is at the half of the distance between Fz and F8. In the study, we targeted F3 for the left dlPFC and F4 for the right dlPFC.

###### **rTMS Parameters.** Active or sham rTMS was performed with a figure-8 coil (A/P Cool-B65) and a MagPro X100 stimulator with MagOption (MagVenture, Denmark). The TMS device was configured to biphasic pulses with the electric field current flowing in the normal direction (AP-PA). Stimulation was delivered over the left or right dlPFC (depending on randomization), defined according to the 10-20 system [73] (F3 and F4, respectively). See supplementary Fig. 1 for details. These procedures allowed for online monitoring and adjustment of TMS coil position throughout the session to ensure proper targeting. rTMS was performed with standard FDA-approved parameters [74,75]. To determine rMT, electrodes (Neuroline 720, Ambu) were placed in a belly-tendon montage on the participant’s opposite from the site of stimulation hand to record activity in the first dorsal interosseous (FDI) muscle, and motor evoked potentials (MEP) were recorded in Brainsight. The motor hot spot was defined as the position over the left/right motor cortex that elicited the largest MEP, and rMT was then defined as the TMS pulse intensity that induced, on average, a MEP of 50 µV amplitude, using a maximum likelihood estimator (TMS Motor Threshold Assessment Tool, MTAT 2.0, clinicalresearcher.org /software.html). For participants who expressed sensitivity to the initial TMS pulses, a ‘ramping up’ procedure was implemented in which the first 2-10 trains were delivered at 80% rMT intensity and increased by 5-10% during the habituation period as tolerated until target intensity was reached.

Each TMS session was conducted by the principal investigator (PI; first author) with the assistance of a TMS technician. The PI led the participant through the session, decided on dose adjustments and course of action for any protocol deviations, and stayed blinded to the condition. The technician was not blind to the assigned condition and prepared the coil but did not influenced the course of the session in any way.

Sham stimulation was delivered using the opposite face of the same A/P coil, which included a magnetic shield to greatly attenuate the induced field. When positioned in the sham configuration, the coil also produced electrical current through two electrodes placed approximately one centimeter apart near the participant’s hairline on the side of the stimulation. This electrical current was matched in intensity to create a somatosensation similar to the active stimulation, thereby creating a reliable sensory blinding to the two conditions. The electrodes were put near the hairline for all participants and were only activated for sham participants. Therefore, the presence of the sham electrodes, the scalp stimulation, the sounds and the coil position over the identified target, and the pre-determination of the motor threshold were all elements of the neurostimulation procedure that were identical between the treatment conditions. The difference was the presence or absence of HF rTMS.

**Combined rTMS-CR Intervention.** First, a 600 s habituation period was performed when participants received active or sham rTMS alone while listening to white noise via headphones. Participants were not instructed to think of anything in particular during this time. Then, the intervention proceeded as follows: (1) participants sat quietly for a 300 s baseline while listening to white noise, (2) participants were instructed to imagine as vividly as possible one of the stressful experiences constructed at intake, (3) the stressor was heard via headphones followed by silence when participants were instructed to continue to imagine the stressful situation (120 s total), and (4) instructions in reducing distress using CR followed. The rTMS began within 10 s after the CR instructions appeared. Reminders of reframing and distancing appeared in random order 180 s and 360 s post-stimulation onset. White noise was played throughout when instructions were not presented. After 600 s, there was a break, followed by a second and third administration of the stressor task using the procedures outlined above, but with a different personalized stressor recording each time presented in a randomized order. After each baseline, stress induction, and regulation period, the participant was asked to rate subjective units of distress (SUDS; 0-9 scale). At the end of the intervention, manipulation check questions ensuring compliance with the instructions and asking participants to guess the blinded condition assignment were administered.

**Design considerations.** Thus, the intervention session included behavioral training, ten minutes of neurostimulation alone to reduce novelty and habituate the participant to the experience, and subsequent administration of rTMS concurrent with engagement in cognitive restructuring during three different regulation periods. There were several possible models to combine neurostimulation and skills training that included interleaved, sequential, or concurrent approaches. Different combination approaches have been more thoroughly examined in the field of neurology [76] where concurrent interventions were slightly favored. Performance optimization studies also favor concurrent designs [77-79]. Therefore, we opted for a concurrent administration of the behavioral and neurostimulation interventions. The mechanistic hypothesis is that both interventions are targeted towards the same neural circuit, and, concurrent administration will enhance long term potentiation and therefore learning [77,80].

We also made the decision to anchor neurostimulation administration to the end of the emotion induction. This decision was guided based on data from the substance abuse and eating disorder literature showing that priming with the problem behavior prior to rTMS enhances the effectiveness of rTMS [81,82]. Experimental findings also show that rTMS facilitates the detection of non-primed targets following priming [83]. Therefore, inducing distress prior to rTMS should enhance the regulatory effect of rTMS.

We decided to divide the concurrent intervention into 3 epochs. On the one hand, we wanted to replicate depression treatment protocols where 40 minutes of neurostimulation is administered. Nevertheless, engagement in autobiographical emotion regulation for 40 minutes was not feasible and receiving rTMS during emotional induction ran the risk of dampening the emotional experience [14]. Thus a design that used continuous stimulation would have introduced unwanted confounds. Breaks between rTMS administration do not reduce effectiveness, but rather enhance facilitatory after-effects [84]. Therefore, we broke the neurostimulation administration in 10 minute epochs concurrent with regulation of three different autobiographical stressors.

###### **Physiological Measurements.** Psychophysiological measurements were collected continuously during the intervention using the BIOPAC MP-150 recording system (Goleta, CA). Electrodes recording heart rate (HR) were placed on the participant’s wrist, ankle, and fingers. Amplified analog data recorded with a 200 Hz sampling rate were converted to digital recording and filtered using BIOPAC’s AcqKnowledge 4.1 software.

##### Ambulatory Assessment and Follow-Up Visit

Following the intervention participants received 8 calls/day for 7 days, starting the day after the intervention, at pseudo-random times to examine SUDS and use of CR. On each call they were asked if they had used CR since the previous call (yes/no) and what was their current level of distress (0- no distress at all to 9- extreme distress). At the end of the week the battery of self-reports from intake was administered again via an online link.

Participants returned to the research office a month later to complete a stressor task without neurostimulation, an acceptability and feasibility interview, and the battery of self-reports. The stressor task included measuring HR while the participant stood still for a 300-s baseline, underwent a 120-s stress induction using a fourth stressor, and engaged in prompted CR for 300 s. Physiological data collection and processing was identical to the intervention procedures. SUDS were rated after each experimental task. After the exit interview, the blind was broken and the experimenter and the subject were debriefed about the study.

**Measures**

***Diagnostic Assessment***

The SCID-5 [26] and SCID-PD [27] were used to assess DSM-5 disorders. Participants were led through both structured interviews by either the first author (76.4% of cases) or one of three trained diagnostic assessors under the supervision of the first author. In the cases where the first author did not conduct the interview, she reviewed in detail with the assessor the questions asked to confirm diagnostic profile. In case of disagreement, she reassessed the disorder at the next visit.

***Self-reports***

**Emotion Regulation Questionnaire (ERQ).** The ERQ [30] is a 10‐item self‐report inventory that assesses the routine use of two cognitive emotion regulation strategies: expressive suppression and cognitive reappraisal. The items use a 7‐point Likert scale with responses ranging from one (*strongly disagree*) to 7 (*strongly agree*). In our study, Cronbach alpha at intake for ERQ Reappraisal was .87. The correlation between phone screen and intake reappraisal scores for 98 participants was .57. ERQ suppression scale scores were collected but not examined for the current study.

**Difficulties in Emotion Regulation Scale (DERS).** The DERS [85] is a 36-item self-report measure of individuals’ typical levels of emotion dysregulation across six domains. Participants respond on a Likert scale ranging from 1 (*almost never*) to 5 (*almost always*), and the total sum score indicates high dysregulation. In the present study, Cronbach’s alpha for the total score at intake was .89. The correlation between phone screen and intake total DERS scores for 93 participants was .66.

**Outcome Questionnaire-45 (OQ-45).** The OQ-45 [86] is a 45-item self-report measure used to track severity of psychopathology throughout treatment. It consists of subscales that identify three types of problems that lead to general stress: psychological symptoms, interpersonal conflicts, and problems with social roles [87]. Items are rated on a Likert scale ranging from 0 (*never*) to 4 (*almost always*). At pretreatment, Cronbach’s alpha for the total score was .83.

**The Work and Social Adjustment Scale (WSAS).** The WSAS [88] is a 5-item self-report inventory examining the functional impairment attributable to an identified problem. In this study, we asked participants to rate the level of impairment that was related to their emotional dysregulation (e.g., “Because of my difficulties managing emotions, my ability to work is impaired”). These questions are rated on a 9-point Likert scale (0 = my problem does not affect this at all; 8 = my problem affects this very seriously) A total WSAS score above 10 suggests clinical levels of functional impairment. At pretreatment, Cronbach’s alpha for the total score was .71.

**Subjective Units of Distress Scale (SUDS).** During the experimental sessions and ambulatory assessments we asked participants to rate their current distress on a scale from 0 – no distress to 9 – extreme distress [89].

**Manipulation Check.** After each baseline and regulation period we examined dissociation during that segment using a 4-item scale [90]. At the end of the intervention and at the 1-week and 1-month follow-up assessments, participants rated their confidence in the assigned condition to which they were kept blind (1 = “I am certain I received sham stimulation” to 9 = “I am certain I received active stimulation”). We also asked after the intervention for participants to give their best guess whether they received real or sham neurostimulation (forced- choice question).

**Tolerability Questionnaire.** Before and after the intervention session, participants rated on a scale from 0–3 (absent, mild, moderate, severe) the intensity of their headache, neck pain, scalp pain, seizure (as observed by technician), hearing impairment and any other side effect that they might have experienced from the TMS treatment.

**Exit Interview.** We refined a previously developed in-house interview [91] to examine feasibility and acceptability as directly relevant to the study and to collect participant feedback (available upon request). The interview was administered at the 1-month follow up, and it included open-ended questions about the overall experience, positives, and issues with the current treatment, as well as Likert-type questions about feasibility of the intervention (e.g., difficulty with limiting movement, level of comfort, ability to concentrate given the TMS noise, distress about the procedures, ease to hear and understand clinician, connection with clinician, and session engagement), acceptability (of session length, skills training, TMS procedures, personalized stressors use, ambulatory phone assessment) and overall satisfaction (i.e., likelihood to recommend to someone else). Feasibility and acceptability questions were rated on a scale from 0 (not at all) to 9 (extremely) and averaged to compute an overall feasibility and acceptability score. Satisfaction was rated on a 0 (low) to 100 (high) continuous scale.

***Psychophysiological Measures***

Raw ECG data were visually inspected and artifacts cleaned prior to calculation of HF-HRV following established guidelines [92,93]. The AcqKnowledge software tool detects R-waves in the cleaned ECG to create a series of inter-beat intervals that is converted to a continuous, time-domain representation of HR using cubic-spline interpolation, which is resampled at 8 Hz. Spectral analysis yields summary report values for HRV frequency bands. Each session period (e.g., baseline, habituation, stressor, regulation) was divided into 120 s bins, and HF-HRV was extracted from each bin. The bin size was chosen because the stressor induction portion was 120s long, and segments were intended to be equivalent in length across the experiment. Therefore, two HF-HRV values for each baseline, one for each stressor, and five values for each regulation period and habituation were computed. HF-HRV was intended to be a primary outcome measure but was added later to clinicaltrials.gov record as primary because of an administrative omission at the beginning.

###### The ‘Find Rate’ function of AcqKnowledge was used to transform the ECG signal into beats per minute or heart rate (HR), using a moving average with a window of 15 s. For each baseline, HR was averaged from the last 240 of the total 300 s. We excluded the first 60 s from each baseline because often during this time participants were still settling into their chair and task and therefore, we considered the first minute not a true representation of physiological baseline. In cases where there was a clear spike in HR (e.g., because the participant coughed, talked, moved abruptly, etc), the average baseline HR was calculated from the maximum time available excluding any amount of time with disruptions.

###### **Regulation Duration.** Time to return to one’s heart rate baseline (regulation duration) was defined for each regulation period as the amount of time it took from the beginning of regulation for the continuously monitored HR to reach a value that was lower or equal to the average baseline HR. If the person started the regulation period below the average baseline, we coded the return to baseline as “never stressed” and did not include it in analyses. If the participant never returned to baseline in the 600 s allocated to the regulation period, we set the return to baseline time to be 600. This data was analyzed with mixed models ANOVA which requires covarying the baseline measurement of the outcome. To create a baseline value for regulation duration, we measured the time it took during the habituation period for the person to return to HR baseline after increased arousal induced by neurostimulation. If the person was not above HR baseline at the beginning of neurostimulation, this covariate was set to 0. If the person never returned to HR baseline during habituation, the covariate was set to 600. At follow up, regulation duration was defined identically. Because only one measurement of regulation duration was achieved, a simpler ANOVA model was planned and a baseline value for this outcome was not needed. An additional difference was that the maximum value the variable could take was 300 s.

**Statistical Analyses**

All analyses compared the effect of sham to active rTMS over the left and right dlPFC. Preliminary analyses including analysis of variance (ANOVA) and chi square tests were conducted to assess demographic differences between groups. We also examined any differences between groups on randomization variables (gender, use of medication) and other potential confounding variables like dissociation during regulation, or arousal induced by rTMS alone. Significant differences were included in subsequent analyses as covariates. Planned covariates included baseline measurements of the outcomes. Analyses were conducted using SPSS version 25.0.

Mixed-effects hierarchical linear models (MMANOVA) with analytically determined covariance structures were used to analyze the repeated measures data [94]. All MMANOVA models used a restricted estimated maximum likelihood model to account for missing data [95] (i.e., cases with missing data were not discarded, but slopes for each participant were computed with the data available). Estimated marginal means (EMMs) were compared using LSD corrections for significant main and interaction effects. Effect sizes for these models were computed using Feingold’s formula [96] and interpreted using Cohen’s [97] specifications.

To test immediate effects of the intervention, we conducted three analyses examining HF-HRV, regulation duration, and SUDS. The treatment condition (active left, active right, or sham), the experimental condition (regulation 1, 2, and 3), the time within each experimental condition (coded 0-4 for each 120 s segment within that period) were used to predict HF-HRV. Baseline HF-HRV was measured at the beginning of the experiment (session baseline) and right before each autobiographical stressor presentation (pre-stimulus baseline). Because active rTMS may have cumulative effects [98] that could influence the pre-stimulus baselines we included both session baseline and pre-stimulus baseline as covariates in the analyses. Because the two HF-HRV values extracted from each baseline (one from the last 120 s, one from the middle 120s) were very highly correlated (*r*s ranging from .93 to .98, *p*s < .001), we only included one of the values for each session and pre-stimulus baseline, corresponding to the last 120 s of the baseline. The SPSS syntax for the HF-HRV analysis was:

MIXED

HF-HRV BY treatment_condition experimental_condition time WITH sessionBaselineHRV prestimulusBaselineHRV

/CRITERIA = CIN(95) MXITER(100) MXSTEP(5) SCORING(1) SINGULAR(0.000000000001) HCONVERGE(0, ABSOLUTE) LCONVERGE(0, ABSOLUTE) PCONVERGE(0.000001, ABSOLUTE)

/FIXED = treatment_condition experimental_condition time sessionBaselineHRV prestimulusBaselineHRV treatment_condition *Time | SSTYPE(3)

/METHOD = REML

/PRINT = SOLUTION TESTCOV R

/REPEATED = experimental_condition time | SUBJECT(SubjectID) COVCONDITION(un)

/EMMEANS = TABLES(treatment_condition) COMPARE ADJ(LSD)

/EMMEANS = TABLES (treatment_condition*time) COMPARE (treatment_condition).

A MMANOVA examining regulation duration during each regulation period was also conducted using treatment condition, experimental condition, and return to emotional baseline during habituation as predictors. SPSS syntax for this analysis was:

MIXED

Regulation_duration BY treatment_condition experimental_condition WITH regulation_duration_baseline

/CRITERIA = CIN(95) MXITER(100) MXSTEP(5) SCORING(1) SINGULAR(0.000000000001) HCONVERGE(0, ABSOLUTE) LCONVERGE(0, ABSOLUTE) PCONVERGE(0.000001, ABSOLUTE)

/FIXED = treatment_condition experimental_condition regulation_duration_baseline | SSTYPE(3)

/METHOD = REML

/PRINT = SOLUTION TESTCOV R

/REPEATED = experimental_condition | SUBJECT(SubjectID) COVCONDITION(AR1)

/EMMEANS = TABLES(treatment_condition) COMPARE ADJ(LSD)

SUDS data were collected and analyzed with two main questions in mind: (a) were the procedures successful in getting participants to feel increased distress after the autobiographical stressors presentation and decreased distress after regulation, and (b) did participants experience lower distress after rTMS enhanced CR when compared to CR alone. Therefore, all SUDS values collected after each pre-stimulus baseline, stressor, and regulation period during the combined intervention were included in the analysis. SUDS after habituation were covaried as the closest ‘baseline’ measurement before the intervention began. Treatment condition, experimental period (post-baseline, post-stressor, post-regulation), and experimental condition (regulation 1, regulation 2, regulation 3) were used to predict SUDS throughout the experiment. SPSS syntax for this analysis was:

MIXED

SUDS BY treatment_condition experimental_condition experimental_period WITH	baselineSUDS

/CRITERIA = CIN(95) MXITER(100) MXSTEP(5) SCORING(1) SINGULAR(0.000000000001) HCONVERGE(0, ABSOLUTE) LCONVERGE(0, ABSOLUTE) PCONVERGE(0.000001, ABSOLUTE)

/FIXED = treatment_condition experimental_condition experimental_period baselineSUDS treatment_condition* experimental_period | SSTYPE(3)

/METHOD = REML

/PRINT = SOLUTION TESTCOV R

/REPEATED = experimental_period	| SUBJECT(SubjectID) COVCONDITION(vc)

/EMMEANS = TABLES(treatment_condition) COMPARE ADJ(LSD)

/EMMEANS = TABLES (treatment_condition* experimental_period) compare (treatment_condition).

Two generalized estimated equations models (GEE) [99] using ordinal logistic models and an independent covariance structure examined differences between treatment conditions in side effects.

To test near-term effects of the TMS-CR intervention a hierarchical linear models (HLM) [100] was used to examine condition differences in SUDS during the ambulatory assessment using the SUDS rating at the beginning of the intervention day (before any procedures were conducted) as baseline. To examine use of CR (indicated as yes/no on each call) during the ambulatory week, we utilized a GEE binary logistic model (logit link, unstructured working correlation with robust estimators) [101,102]. Intake ERQ reappraisal was included as a co-variate. Call days were considered a continuous time variable, and call number within the day a categorical one. SPSS syntax for both analyses is listed below.

GENLIN use_of_CR_skill (REFERENCE=FIRST) BY treatment_condition callNumber (ORDER=ASCENDING) WITH ERQ_reappraisal_baseline callDay

/MODEL treatmentcondition callDay callNumber ERQ_reappraisal_baseline INTERCEPT=YES

DISTRIBUTION=BINOMIAL LINK=LOGIT

/CRITERIA METHOD=FISHER(1) SCALE=1 MAXITERATIONS=100 MAXSTEPHALVING=5 PCONVERGE=1E-006(ABSOLUTE) SINGULAR=1E-012 ANALYSISTYPE=3(WALD) CILEVEL=95 LIKELIHOOD=FULL

/EMMEANS SCALE=ORIGINAL

/EMMEANS TABLES=treatment_condition SCALE=ORIGINAL COMPARE=treatment_condition CONTRAST=SIMPLE(2) PADJUST=LSD

/REPEATED SUBJECT=SubjectID WITHINSUBJECT=callDay*callNumber SORT=YES CORRTYPE=UNSTRUCTURED

ADJUSTCORR=YES COVB=ROBUST MAXITERATIONS=1000000000 PCONVERGE=1e-006(ABSOLUTE) UPDATECORR=1

/MISSING CLASSMISSING=EXCLUDE

/PRINT CPS DESCRIPTIVES MODELINFO FIT SUMMARY SOLUTION.

MIXED

SUDS BY Subjectid treatment_condition callNumber WITH callDay SUDS_beforeIntervention

/CRITERIA = CIN(95) MXITER(1000000000) MXSTEP(5) SCORING(1)

SINGULAR(0.000000000001) HCONVERGE(0, ABSOLUTE) LCONVERGE(0, ABSOLUTE)

PCONVERGE(0.000001, ABSOLUTE)

/FIXED = condition callDay callNumber SUDS_beforeIntervention	| SSTYPE(3)

/METHOD = REML

/PRINT = COVB SOLUTION TESTCOV

/RANDOM INTERCEPT callDay callNumber| SUBJECT(SubjectID) COVTYPE(id)

/EMMEANS = TABLES(treatment_condition) COMPARE ADJ(LSD).

To test the long-term effects of the intervention, six MMANOVA models were conducted: four examining between-condition differences at the 1-week and 1-month follow-up assessments in ERQ, DERS, OQ-45, and WSAS; and two examining HF-HRV and SUDS during the follow-up stressor task. An ANCOVA examined regulation duration differences between conditions at follow up. For the longitudinal self-reports (DERS, OQ-45, WSAS, and ERQ Reappraisal), intake measurements were used as covariates in the analyses. HF-HRV was extracted from the first 120 s period and the next 120 s period out of the total 300 s regulation period. The treatment condition and baseline HF-HRV (extracted from the last 2-min of baseline) were added as covariates. Treatment condition, time period in the follow-up stressor task (post-baseline, post stressor, post regulation), SUDS baseline (collected at the beginning of the 1-month follow-up day) were used to predict SUDS at follow up. In addition, we conducted a univariate general linear model examining differences between conditions in the time it took to return to baseline during the regulation period at follow up. No covariates were included. SPSS syntax is passed below (OUTCOME can be either DERS, ERQ_Reappraisal, WSAS, or OQ-45 measured at the one week and one month follow up; OUTCOME_baseline is the value of the self report at intake).

MIXED

OUTCOME BY condition time WITH OUTCOME_baseline

/CRITERIA = CIN(95) MXITER(100) MXSTEP(5) SCORING(1)

SINGULAR(0.000000000001) HCONVERGE(0, ABSOLUTE) LCONVERGE(0, ABSOLUTE)

PCONVERGE(0.000001, ABSOLUTE)

/FIXED = treatment_condition time OUTCOME_baseline treatment_condition*time | SSTYPE(3)

/METHOD = REML

/PRINT = SOLUTION TESTCOV R

/REPEATED =	Time | SUBJECT(SubjectID) COVCONDITION(un)

/EMMEANS = TABLES(treatment_condition) COMPARE ADJ(LSD)

/EMMEANS = TABLES (treatment_condition*time) COMPARE (CONDITION).

MIXED

HF-HRV_follow_up BY treatment_condition Time WITH baselineHRV_follow_up

/CRITERIA = CIN(95) MXITER(100) MXSTEP(5) SCORING(1)

SINGULAR(0.000000000001) HCONVERGE(0, ABSOLUTE) LCONVERGE(0, ABSOLUTE)

PCONVERGE(0.000001, ABSOLUTE)

/FIXED = treatment_condition Time baselineHRV_follow_up	| SSTYPE(3)

/METHOD = REML

/PRINT = SOLUTION TESTCOV R

/REPEATED =	Time | SUBJECT(SubjectID) COVCONDITION(cs)

/EMMEANS = TABLES(treatment_condition) COMPARE ADJ(LSD).

UNIANOVA regulation_period_followUP BY treatment_condition

/CONTRAST(treatment_condition)=Simple

/METHOD=SSTYPE(3)

/INTERCEPT=INCLUDE

/POSTHOC= treatment_condition (LSD)

/PLOT=PROFILE(treatment_condition) TYPE=BAR ERRORBAR=NO MEANREFERENCE=NO

/EMMEANS=TABLES(treatment_condition) COMPARE ADJ(LSD)

/PRINT ETASQ TEST(LMATRIX) DESCRIPTIVE PARAMETER HOMOGENEITY OPOWER

/PLOT=SPREADLEVEL

/CRITERIA=ALPHA(.05)

/DESIGN= treatment_condition.

MIXED

SUDS_follow_up BY treatment_condition Time WITH SUDS_follow_up_baseline

/CRITERIA = CIN(95) MXITER(100) MXSTEP(5) SCORING(1)

SINGULAR(0.000000000001) HCONVERGE(0, ABSOLUTE) LCONVERGE(0, ABSOLUTE)

PCONVERGE(0.000001, ABSOLUTE)

/FIXED = treatment_condition Time SUDS_follow_up_baseline treatment_condition *time | SSTYPE(3)

/METHOD = REML

/PRINT = SOLUTION TESTCOV R

/REPEATED =	Time | SUBJECT(SubjectID) COVCONDITION(vc)

/EMMEANS = TABLES(treatment_condition) COMPARE ADJ(LSD)

/EMMEANS = TABLES (treatment_condition *time) COMPARE (treatment_condition).

In cases where significant differences between conditions could not be found, we employed equivalence analyses to examine whether the lack of differences was due to equivalence between conditions. Given the novel intervention and approach, we selected a conservative symmetric margin of .5 as recommended by others [103]. In order to examine equivalence for repeated measures analyses, we used an online calculator (<https://www.psychometrica.de/effect_size.html>; *effect size estimates in repeated measures designs*) and inputted means, standard deviations, and pre-post correlations for measures of interest in order to determine the 90% confidence interval (CI) for the effect size observed. For measures that were not repeated (e.g., return to baseline at follow up), the *comparisons of groups with different sample sizes* from the same calculator was utilized to establish the 90% CI for the effect size observed. This CI corresponds to the two one-sided procedure (TOST)[104] and, when compared to the interval defined by the Smallest Effect Size of Interest (SESOI CI: -0.5 – 0.5), can indicate whether the conditions were equivalent in their outcomes or not. If the bounds of the computed 90% CI were within the SESOI CI, then the conditions were equivalent. If the bounds of the computed 90% CI were outside of the SESOI CI, then the hypothesis that the conditions were equivalent was rejected.

**Supplemental Results**

**Missing data.** One participant had incomplete HF-HRV data for the last regulation period during intervention because the participant wanted to finish early. One participant did not complete the 1-week follow-up battery of self-reports, and one participant missed filing in the WSAS at intake. Two people completed their one-month follow-up assessment online and therefore do not have HF-HRV and regulation duration data for this assessment point. Two participants were lost to contact by the one-month follow-up and did not complete self-reports.

Four participants could not tolerate rTMS above MT. Two of the participants who could not tolerate TMS dropped right after the habituation procedure and, therefore, do not have data for the remainder of the study. One participant continued the intervention at below MT stimulation and completed the study. One participant stopped after the first stressor and regulation period but continued with the study assessments.

Of the 2133 calls placed during the ambulatory assessment, participants did not answer 735 calls, provided invalid data on 5 calls, and were not called because of administrative problems in 4 instances (29.50% of calls). Three participants missed a full day of calls (i.e., did not answer any call), and one participant missed 2 full days of calls.

#### Preliminary Analyses. There were no significant differences between conditions in age, gender distribution, marital status, sexual orientation, ethnicity and race, income, self-reported use of reappraisal, presence of a depressive disorder, use of psychotropic medication, or number of current or lifetime diagnoses met (ps > .05). Therefore, our randomization procedures were successful, and none of these variables were included in the outcome analyses as covariates.

Self-reported dissociation during the experimental tasks was minimal (M_Dissociation_:0.10, SD = 0.41) and, therefore, was not included as a covariate. To examine if rTMS alone affected HF-HRV or SUDS, we conducted MMANOVA analyses using the data collected during the habituation period (i.e., when participants were administered rTMS or sham alone for 10 minutes without any other instructions) and controlling for baseline. There was no significant difference between treatment conditions in HF-HRV or SUDS (*p*s > 0.64) during habituation (M_LG_HF-HRV_ = 1.90; SD = 0.51; M_Sqrt_SUDS_ = 1.25, SD = 0.81). Therefore, habituation HF-HRV and SUDs were not included as covariates, and SDs from these measurements were used to compute effect sizes.

**Supplemental Discussion Points**

With regards to which target was optimal to enhance emotion regulation, our results were mixed. Both left and right targeting increased HF-HRV significantly above sham stimulation. Right rTMS was significantly better than sham in terms of reducing the regulation duration, while left rTMS was only marginally significant. During the week after the intervention only left rTMS showed significant improvements above sham in skill acquisition and reduced distress. The literature is also inconsistent, with benefits to emotion regulation being shown in studies that employ either left or right rTMS [11,12,105]. Neuroscientific meta-analyses point to increases in both right and left dlPFC recruitment for downregulation when compared to upregulation of emotion [67,106]. In addition, the left- versus right-recruitment may depend on the tactic employed[63]. Therefore, response to left or right dlPFC stimulation may depend on the tactic that was easier for each participant to apply. Taken together, these studies and our findings suggest that engaging either the right or the left dlPFC with emotion regulation or neurostimulation is effective in reducing emotional arousal. Furthermore, excitatory neurostimulation administered over the left or right dlPFC may appear to have different effects depending on the outcome measure administered. Further work is needed to better understand differences between left and right rTMS.

### **References**

1.	DuPont RL, Rice DP, Miller LS, Shiraki SS, Rowland CR, Harwood HJ. Economic costs of anxiety disorders. Anxiety. 1996;2(4):167-72.

2.	Greenberg PE, Kessler RC, Birnbaum HG, Leong SA, Lowe SW, Berglund PA, et al. The economic burden of depression in the United States: how did it change between 1990 and 2000? J Clin Psych. 2003;64(12):1465-75.

3.	Lépine J-P, Briley M. The increasing burden of depression. Neuropsychiatr Dis Treat. 2011;7(Suppl 1):3.

4.	Schmitz N, Kruse J. The relationship between mental disorders and medical service utilization in a representative community sample. Soc Psychiatry Psychiatr Epidemiol. 2002;37(8):380-6.

5.	Carta MG, Sancassiani F, Pippia V, Bhat KM, Sardu C, Meloni L. Alexithymia is associated with delayed treatment seeking in acute myocardial infarction. Psychother Psychosom. 2013;82(3):190-2.

6.	Ciarrochi JV, Deane FP. Emotional competence and willingness to seek help from professional and nonprofessional sources. Br J Guidance & Counseling. 2001;29:233-46.

7.	Vogel DL, Wade NG, Hackler AH. Emotional Expression and the Decision to Seek Therapy: The Mediating Roles of the Anticipated Benefits and Risks. J Soc Clin Psycol. 2008;27(3):254-78.

8.	Hasani N, Oghabian M, Arbabi M, Hafizi S, Gholami S, editors. Evaluation of fMRI analysis to study treatment effect of TMS in borderline personality disorder. World Congress on Medical Physics and Biomedical Engineering May 26-31, 2012, Beijing, China; 2013: Springer.

9.	Neacsiu AD, Lisanby SH. Magnetic stimulation for depression: Subconvulsive and convulsive approaches. Neuromodulation in Psychiatry. 2015:155-80.

10.	Balconi M, Ferrari C. rTMS stimulation on left dlpfc affects emotional cue retrieval as a function of anxiety level and gender. Depress Anxiety. 2012;29(11):976-82.

11.	De Raedt R, Leyman L, Baeken C, Van Schuerbeek P, Luypaert R, Vanderhasselt M-A, et al. Neurocognitive effects of HF-rTMS over the dorsolateral prefrontal cortex on the attentional processing of emotional information in healthy women: An event-related fMRI study. Biol Psychology. 2010;85(3):487-95.

12.	Feeser M, Prehn K, Kazzer P, Mungee A, Bajbouj M. Transcranial Direct Current Stimulation Enhances Cognitive Control During Emotion Regulation. Brain Stimulation. 2014;7(1):105-12.

13.	Leyman L, De Raedt R, Vanderhasselt MA, Baeken C. Influence of high-frequency repetitive transcranial magnetic stimulation over the dorsolateral prefrontal cortex on the inhibition of emotional information in healthy volunteers. Psychol Med. 2009;39(6):1019-28.

14.	Remue J, Vanderhasselt M-A, Baeken C, Rossi V, Tullo J, De Raedt R. The effect of a single HF-rTMS session over the left DLPFC on the physiological stress response as measured by heart rate variability. Neuropsychology. 2016;30(6):756.

15.	Thayer JF, Åhs F, Fredrikson M, Sollers III JJ, Wager TD. A meta-analysis of heart rate variability and neuroimaging studies: implications for heart rate variability as a marker of stress and health. Neurosci Biobehav Rev. 2012;36(2):747-56.

16.	Lane RD, McRae K, Reiman EM, Chen K, Ahern GL, Thayer JF. Neural correlates of heart rate variability during emotion. NeuroImage. 2009;44(1):213-22.

17.	Appelhans BM, Luecken LJ. Heart rate variability as an index of regulated emotional responding. Rev Gen Psychol. 2006;10(3):229-40.

18.	Malik M. Heart rate variability: Standards of measurement, physiological interpretation, and clinical use: Task force of the European Society of Cardiology and the North American Society for Pacing and Electrophysiology. Annals of Noninvasive Electrocardiology. 1996 Apr;1(2):151-81.

19.	Geisler F, Kubiak T. Heart rate variability predicts self‐control in goal pursuit. Eur J Personality. 2009;23(8):623-33.

20.	Butler EA, Wilhelm FH, Gross JJ. Respiratory sinus arrhythmia, emotion, and emotion regulation during social interaction. Psychophysiology. 2006;43(6):612-22.

21.	Demaree HA, Robinson JL, Everhart DE, Schmeichel BJ. Resting RSA is associated with natural and self-regulated responses to negative emotional stimuli. Brain Cogn. 2004;56(1):14-23.

22.	Elliot AJ, Payen V, Brisswalter J, Cury F, Thayer JF. A subtle threat cue, heart rate variability, and cognitive performance. Psychophysiology. 2011;48(10):1340-5.

23.	Fenton-O'Creevy M, Lins JT, Vohra S, Richards DW, Davies G, Schaaff K. Emotion regulation and trader expertise: Heart rate variability on the trading floor. J Neurosci Psychol Econ. 2012;5(4):227-37.

24.	Cunningham DA, Varnerin N, Machado A, Bonnett C, Janini D, Roelle S, et al. Stimulation targeting higher motor areas in stroke rehabilitation: A proof-of-concept, randomized, double-blinded placebo-controlled study of effectiveness and underlying mechanisms. Restor Neurol Neurosci. 2015;33(6):911-26.

25.	Bentwich J, Dobronevsky E, Aichenbaum S, Shorer R, Peretz R, Khaigrekht M, et al. Beneficial effect of repetitive transcranial magnetic stimulation combined with cognitive training for the treatment of Alzheimer’s disease: a proof of concept study. J Neural Transm. 2011;118(3):463-71.

26.	First MB, Williams JB, Karg RL. Structured Clinical Interview for DSM-5 Disorders (Research Version: SCID-5-RV). Washington, DC: American Psychiatric Publishing; 2015.

27.	First MB, Williams JB, Benjamin LS, Spitzer RL. SCID-5-PD: Structured clinical interview for DSM-5® personality disorders: American Psychiatric Association Publishing; 2016.

28.	Fernandez KC, Jazaieri H, Gross JJ. Emotion Regulation: A Transdiagnostic Perspective on a New RDoC Domain. Cognit Ther Res. 2016;40(3):426-40.

29.	Insel T, Cuthbert B, Garvey M, Heinssen R, Pine DS, Quinn K, et al. Research Domain Criteria (RDoC): Toward a New Classification Framework for Research on Mental Disorders. Am J Psych. 2010;167(7):748-51.

30.	Gross JJ, John OP. Individual differences in two emotion regulation processes: Implications for affect, relationships, and well-being. J Pers Soc Psychol. 2003;85(2):348-62.

31.	Aldao A, Nolen-Hoeksema S, Schweizer S. Emotion-regulation strategies across psychopathology: A meta-analytic review. ClinPsychol Rev. 2010;30(2):217-37.

32.	Amstadter A. Emotion regulation and anxiety disorders. Journal of anxiety disorders. 2008;22(2):211-21.

33.	Melka SE, Lancaster SL, Bryant AR, Rodriguez BF. Confirmatory factor and measurement invariance analyses of the emotion regulation questionnaire. J Clin Psychol. 2011;67(12):1283-93.

34.	Salsman NL, Linehan MM. An investigation of the relationships among negative affect, difficulties in emotion regulation, and features of borderline personality disorder. J Psychopathol Behav Assessment. 2012;34(2):260-7.

35.	Dennis TA. Interactions between emotion regulation strategies and affective style: Implications for trait anxiety versus depressed mood. Motiv Emot. 2007;31(3):200-7.

36.	Fresco DM, Moore MT, van Dulmen MH, Segal ZV, Ma SH, Teasdale JD, et al. Initial psychometric properties of the experiences questionnaire: validation of a self-report measure of decentering. Behavior Therapy. 2007;38(3):234-46.

37.	Gillanders S, Wild M, Deighan C, Gillanders D. Emotion regulation, affect, psychosocial functioning, and well-being in hemodialysis patients. Am J Kidney Dis. 2008;51(4):651-62.

38.	Matsumoto D, Yoo SH, Nakagawa S. Culture, emotion regulation, and adjustment. J Pers Soc Psychol. 2008;94(6):925-37.

39.	Haga SM, Kraft P, Corby E-K. Emotion regulation: Antecedents and well-being outcomes of cognitive reappraisal and expressive suppression in cross-cultural samples. J Happiness Studies. 2009;10(3):271-91.

40.	Zuurbier LA, Nikolova YS, Åhs F, Hariri AR. Uncinate fasciculus fractional anisotropy correlates with typical use of reappraisal in women but not men. Emotion. 2013;13(3):385.

41.	Vanderhasselt M-A, Baeken C, Van Schuerbeek P, Luypaert R, De Raedt R. Inter-individual differences in the habitual use of cognitive reappraisal and expressive suppression are associated with variations in prefrontal cognitive control for emotional information: an event related fMRI study. Biol Psychology. 2013;92(3):433-9.

42.	Swart M, Kortekaas R, Aleman A. Dealing with feelings: characterization of trait alexithymia on emotion regulation strategies and cognitive-emotional processing. PLoS One. 2009;4(6):e5751.

43.	Li Z, Sang Z, Zhang Z. Expressive suppression and financial risk taking: A mediated moderation model. Personality and Individual Differences. 2015;72:35-40.

44.	Egloff B, Schmukle SC, Burns LR, Schwerdtfeger A. Spontaneous emotion regulation during evaluated speaking tasks: associations with negative affect, anxiety expression, memory, and physiological responding. Emotion. 2006;6(3):356-66.

45.	Orgeta V. Avoiding Threat in Late Adulthood: Testing Two Life Span Theories of Emotion. Experimental Aging Research. 2011;37(4):449-72.

46.	Joormann J, Gotlib IH. Emotion regulation in depression: Relation to cognitive inhibition. Cogn Emot. 2010;24(2):281-98.

47.	Moore SA, Zoellner LA, Mollenholt N. Are expressive suppression and cognitive reappraisal associated with stress-related symptoms? Behav Res Ther. 2008;46(9):993-1000.

48.	Dunn LM. PPVT-revised manual. Circle Pines, MN: American Guidance Service; 1981.

49.	Pitman RK, Orr SP, Forgue DF, de Jong JB, Claiborn JM. PSychophysiologic assessment of posttraumatic stress disorder imagery in vietnam combat veterans. Arch Gen Psychiatry. 1987;44(11):970-5.

50.	Schmahl CG, Elzinga BM, Ebner UW, Simms T, Sanislow C, Vermetten E, et al. Psychophysiological reactivity to traumatic and abandonment scripts in borderline personality and posttraumatic stress disorders: a preliminary report. Psychiatry Res. 2004;126(1):33-42.

51.	Scott NW, McPherson GC, Ramsay CR, Campbell MK. The method of minimization for allocation to clinical trials. A review. Control Clin Trials. 2002;23(6):662-74.

52.	Taves DR. Minimization: a new method of assigning patients to treatment and control groups. Clin Pharmacol Ther. 1974;15(5):443-53.

53.	Linehan M. DBT Skills training manual. 2^nd^ edition. New York, NY: Guilford Publications; 2014.

54.	Beck A, Emery G, Greenberg R. Anxiety Disorders and Phobias: A Cognitive Perspective. New York: Basic Books. 1985.

55.	Shurick AA, Hamilton JR, Harris LT, Roy AK, Gross JJ, Phelps EA. Durable effects of cognitive restructuring on conditioned fear. Emotion. 2012;12(6):1393-7.

56.	Katz L, Epstein S. Constructive thinking and coping with laboratory-induced stress. J Pers Soc Psychol. 1991;61(5):789-800.

57.	Lazarus RS, Folkman S. Stress, appraisal, and coping. Springer publishing company; 1984.

58.	Meichenbaum D. Stress inoculation training. Pergamon; 1985.

59.	Brock TC. Cognitive restructuring and attitude change. The Journal of Abnormal and Social Psychology. 1962;64(4):264-71.

60.	Gross JJ. Antecedent- and response-focused emotion regulation: Divergent consequences for experience, expression, and physiology. J Pers Soc Psychol. 1998;74(1):224-37.

61.	Speisman JC, Lazarus RS, Mordkoff A, Davison L. Experimental reduction of stress based on ego-defense theory. J Abnorm Psychol. 1964;68:367-80.

62.	Beck AT, Dozois DJ. Cognitive therapy: current status and future directions. Annu Rev Med. 2011;62:397-409.

63.	Powers JP, LaBar KS. Regulating emotion through distancing: A taxonomy, neurocognitive model, and supporting meta-analysis. Neurosci Biobehav Rev. 2019;96:155-73.

64.	Ochsner KN, Bunge SA, Gross JJ, Gabrieli JDE. Rethinking feelings: an FMRI study of the cognitive regulation of emotion. J Cogn Neurosci. 2002;14(8):1215-29.

65.	Kendall PC. Coping cat workbook. Workbook Pub; 2006.

66.	Pico-Perez M, Radua J, Steward T, Menchon JM, Soriano-Mas C. Emotion regulation in mood and anxiety disorders: A meta-analysis of fMRI cognitive reappraisal studies. Prog Neuropsychopharmacol Biol Psychiatry. 2017;79(Pt B):96-104.

67.	Ochsner KN, Silvers JA, Buhle JT. Functional imaging studies of emotion regulation: a synthetic review and evolving model of the cognitive control of emotion. Ann N Y Acad Sci. 2012;1251:E1-24.

68.	Kroes MC, Dunsmoor JE, Hakimi M, Oosterwaal S, collaboration NP, Meager MR, et al. Patients with dorsolateral prefrontal cortex lesions are capable of discriminatory threat learning but appear impaired in cognitive regulation of subjective fear. Social cognitive and affective neuroscience. 2019;14(6):601-12.

69.	Gross JJ, Jazaieri H. Emotion, emotion regulation, and psychopathology: An affective science perspective. Clinical Psychological Science. 2014;2(4):387-401.

70.	Neacsiu AD, Bohus M, Linehan MM. Dialectical Behavior Therapy: An Intervention for Emotion Dysregulation. In: Gross, JJ, editor. Handbook of emotion regulation. 2 ed. New York, NY: The Guilford Press; 2013. p. 491-508.

71.	Shalom JG, Aderka IM. A meta-analysis of sudden gains in psychotherapy: outcome and moderators. Clin Psychol Rev. 2020;76:101827.

72.	Anderson EM, Lambert MJ. A survival analysis of clinically significant change in outpatient psychotherapy. J Clin Psychol. 2001;57(7):875-88.

73.	Herwig U, Satrapi P, Schönfeldt-Lecuona C. Using the International 10-20 EEG System for Positioning of Transcranial Magnetic Stimulation. Brain Topography. 2003;16(2):95-9.

74.	Rossi S, Hallett M, Rossini PM, Pascual-Leone A, Group SoTC. Safety, ethical considerations, and application guidelines for the use of transcranial magnetic stimulation in clinical practice and research. Clin Neurophysiol. 2009;120(12):2008-39.

75.	Wassermann EM. Risk and safety of repetitive transcranial magnetic stimulation: report and suggested guidelines from the International Workshop on the Safety of Repetitive Transcranial Magnetic Stimulation, June 5–7, 1996. Electroencephalogr Clin Neurophysiol. 1998;108(1):1-16.

76.	Tsagaris KZ, Labar DR, Edwards DJ. A framework for combining rTMS with behavioral therapy. Front Syst Neurosci. 2016;10:1-8.

77.	Luber BM, Davis S, Bernhardt E, Neacsiu A, Kwapil L, Lisanby SH, et al. Using neuroimaging to individualize TMS treatment for depression: Toward a new paradigm for imaging-guided intervention. NeuroImage. 2017;148:1-7.

78.	Luber B, Steffener J, Tucker A, Habeck C, Peterchev AV, Deng ZD, et al. Extended remediation of sleep deprived-induced working memory deficits using fMRI-guided transcranial magnetic stimulation. Sleep. 2013;36(6):857-71.

79.	Luber B, Stanford AD, Bulow P, Nguyen T, Rakitin BC, Habeck C, et al. Remediation of sleep-deprivation-induced working memory impairment with fMRI-guided transcranial magnetic stimulation. Cerebral cortex. 2008;18(9):2077-85.

80.	De Raedt R, Vanderhasselt M-A, Baeken C. Neurostimulation as an intervention for treatment resistant depression: From research on mechanisms towards targeted neurocognitive strategies. Clin Psychol Rev. 2015;41:61-9.

81.	Van den Eynde F, Claudino AM, Mogg A, Horrell L, Stahl D, Ribeiro W, et al. Repetitive transcranial magnetic stimulation reduces cue-induced food craving in bulimic disorders. Biol Psychiatry. 2010;67(8):793-5.

82.	Amiaz R, Levy D, Vainiger D, Grunhaus L, Zangen A. Repeated high‐frequency transcranial magnetic stimulation over the dorsolateral prefrontal cortex reduces cigarette craving and consumption. Addiction. 2009;104(4):653-60.

83.	Cattaneo Z, Rota F, Vecchi T, Silvanto J. Using state‐dependency of transcranial magnetic stimulation (TMS) to investigate letter selectivity in the left posterior parietal cortex: A comparison of TMS‐priming and TMS‐adaptation paradigms. Eur J Neurosci. 2008;28(9):1924-9.

84.	Rothkegel H, Sommer M, Paulus W. Breaks during 5 Hz rTMS are essential for facilitatory after effects. Clin Neurophysiol. 2010;121(3):426-30.

85.	Gratz KL, Roemer L. Multidimensional Assessment of Emotion Regulation and Dysregulation: Development, Factor Structure, and Initial Validation of the Difficulties in Emotion Regulation Scale. J Psychopathol Behav Assess. 2004;26(1):41-54.

86.	Lambert MJ, Burlingame GM, Umphress V, Hansen NB, Vermeersch DA, Clouse GC, et al. The reliability and validity of the Outcome Questionnaire. Clinical Psychology & Psychotherapy: Intl J Theory Practice. 1996;3(4):249-58.

87.	Wells MG, Burlingame GM, Lambert MJ, Hoag MJ, Hope CA. Conceptualization and measurement of patient change during psychotherapy: Development of the Outcome Questionnaire and Youth Outcome Questionnaire. Psychother Theory Res Prac Train. 1996;33(2):275-283.

88.	Mundt JC, Marks IM, Shear MK, Greist JM. The Work and Social Adjustment Scale: a simple measure of impairment in functioning. Br J Psychiatry. 2002;180(5):461-4.

89.	Wolpe J. The practice of behavior therapy. New York, NY: Pergamon Press; 1969.

90.	Stiglmayr C, Schmahl C, Bremner JD, Bohus M, Ebner-Priemer U. Development and psychometric characteristics of the DSS-4 as a short instrument to assess dissociative experience during neuropsychological experiments. Psychopathology. 2009;42(6):370-4.

91.	Neacsiu AD, Luber BM, Davis SW, Bernhardt E, Strauman TJ, Lisanby SH. On the Concurrent Use of Self-System Therapy and Functional Magnetic Resonance Imaging-Guided Transcranial Magnetic Stimulation as Treatment for Depression. J ECT. 2018;34(4):266-73.

92.	Camm A, Malik M, Bigger J, Breithardt G, Cerutti S, Cohen R, et al. Heart rate variability: standards of measurement, physiological interpretation and clinical use. Task Force of the European Society of Cardiology and the North American Society of Pacing and Electrophysiology. Circulation. 1996;93(5):1043-65.

93.	Berntson GG, Bigger JT, Jr., Eckberg DL, Grossman P, Kaufmann PG, Malik M, et al. Heart rate variability: origins, methods, and interpretive caveats. Psychophysiology. 1997;34(6):623-48.

94.	Molenberghs G, Verbeke G. Linear mixed models for longitudinal data. Springer; 2000.

95.	Gibbons RD, Hedeker D, DuToit S. Advances in analysis of longitudinal data. Annu Rev Clin Psychol. 2010;6:79-107.

96.	Feingold A. Effect sizes for growth-modeling analysis for controlled clinical trials in the same metric as for classical analysis. Psychol Methods. 2009;14(1):43-53.

97.	Cohen J. Statistical power analysis for the behavioral sciences. revised edition. New York, NY: Academic Press; 1977.

98.	Bäumer T, Lange R, Liepert J, Weiller C, Siebner HR, Rothwell JC, et al. Repeated premotor rTMS leads to cumulative plastic changes of motor cortex excitability in humans. Neuroimage. 2003;20(1):550-60.

99.	Liang K-Y, Zeger SL. Longitudinal Data Analysis Using Generalized Linear Models. Biometrika. 1986;73(1):13-22.

100.	Raudenbush SW, Bryk AS. Hierarchical linear models: Applications and data analysis methods. Sage; 2002.

101.	Zeger SL, Liang K-Y. Longitudinal data analysis for discrete and continuous outcomes. Biometrics. 1986:121-30.

102.	Zeger SL, Liang K-Y, Albert PS. Models for longitudinal data: a generalized estimating equation approach. Biometrics. 1988:1049-60.

103.	Norman GR, Sloan JA, Wyrwich KW. Interpretation of changes in health-related quality of life: the remarkable universality of half a standard deviation. Med Care. 2003:582-92.

104.	Lakens D, Scheel AM, Isager PM. Equivalence testing for psychological research: A tutorial. Adv Methods Practices Psychol Sci. 2018;1(2):259-69.

105. Fonzo GA, Goodkind MS, Oathes DJ, Zaiko YV, Harvey M, Peng KK, et al. PTSD psychotherapy outcome predicted by brain activation during emotional reactivity and regulation. Am J Psychiatry. 2017;174(12):1163-74.

106. Kohn N, Eickhoff SB, Scheller M, Laird AR, Fox PT, Habel U. Neural network of cognitive emotion regulation—an ALE meta-analysis and MACM analysis. NeuroImage. 2014;87:345-55.

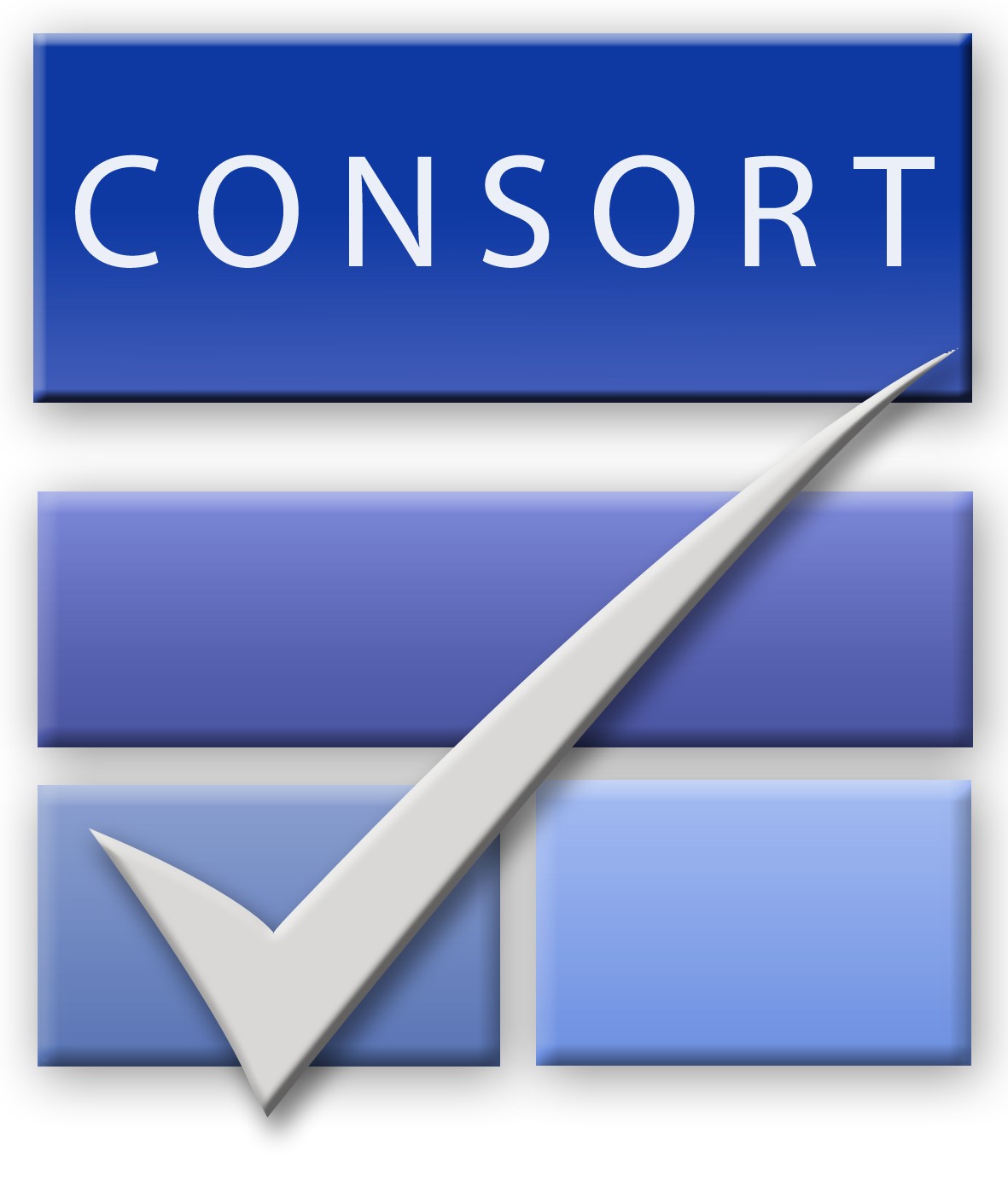
**CONSORT 2010 checklist of information to include when reporting a randomised trial***

| **Section/Topic** | **Item No** | **Checklist item** | **Reported on page No** |
| --- | --- | --- | --- |
| **Title and abstract** | | | |
|  | 1a | Identification as a randomised trial in the title | 1 |
|  | 1b | Structured summary of trial design, methods, results, and conclusions (for specific guidance see CONSORT for abstracts) | 2 |
| **Introduction** | | | |
| Background and objectives | 2a | Scientific background and explanation of rationale | 3 |
|  | 2b | Specific objectives or hypotheses | 4 |
| **Methods** | | | |
| Trial design | 3a | Description of trial design (such as parallel, factorial) including allocation ratio | 4 |
|  | 3b | Important changes to methods after trial commencement (such as eligibility criteria), with reasons | Supplement |
| Participants | 4a | Eligibility criteria for participants | Supplement  CONSORT diagram |
|  | 4b | Settings and locations where the data were collected | Supplement |
| Interventions | 5 | The interventions for each group with sufficient details to allow replication, including how and when they were actually administered | 4-5  Supplement |
| Outcomes | 6a | Completely defined pre-specified primary and secondary outcome measures, including how and when they were assessed | 5-6 |
|  | 6b | Any changes to trial outcomes after the trial commenced, with reasons | Supplement |
| Sample size | 7a | How sample size was determined | 4 |
|  | 7b | When applicable, explanation of any interim analyses and stopping guidelines | n/a |
| Randomisation: |  |  |  |
| Sequence generation | 8a | Method used to generate the random allocation sequence | Supplement |
|  | 8b | Type of randomisation; details of any restriction (such as blocking and block size) | 4, supplement |
| Allocation concealment mechanism | 9 | Mechanism used to implement the random allocation sequence (such as sequentially numbered containers), describing any steps taken to conceal the sequence until interventions were assigned | 4; Supplement |
| Implementation | 10 | Who generated the random allocation sequence, who enrolled participants, and who assigned participants to interventions | Supplement |
| Blinding | 11a | If done, who was blinded after assignment to interventions (for example, participants, care providers, those assessing outcomes) and how | 3; Supplement |
|  | 11b | If relevant, description of the similarity of interventions | Supplement |
| Statistical methods | 12a | Statistical methods used to compare groups for primary and secondary outcomes | 6; Supplement |
|  | 12b | Methods for additional analyses, such as subgroup analyses and adjusted analyses | Supplement |
| **Results** | | | |
| Participant flow (a diagram is strongly recommended) | 13a | For each group, the numbers of participants who were randomly assigned, received intended treatment, and were analysed for the primary outcome | CONSORT flow diagram |
|  | 13b | For each group, losses and exclusions after randomisation, together with reasons | 6, Figure 1 |
| Recruitment | 14a | Dates defining the periods of recruitment and follow-up | 4; Supplement |
|  | 14b | Why the trial ended or was stopped | Supplement |
| Baseline data | 15 | A table showing baseline demographic and clinical characteristics for each group | Table 1 |
| Numbers analysed | 16 | For each group, number of participants (denominator) included in each analysis and whether the analysis was by original assigned groups | CONSORT flow diagram Supplement |
| Outcomes and estimation | 17a | For each primary and secondary outcome, results for each group, and the estimated effect size and its precision (such as 95% confidence interval) | 6-9 |
|  | 17b | For binary outcomes, presentation of both absolute and relative effect sizes is recommended | n/a |
| Ancillary analyses | 18 | Results of any other analyses performed, including subgroup analyses and adjusted analyses, distinguishing pre-specified from exploratory | 6-9; Supplement |
| Harms | 19 | All important harms or unintended effects in each group (for specific guidance see CONSORT for harms) | 7 |
| **Discussion** | | | |
| Limitations | 20 | Trial limitations, addressing sources of potential bias, imprecision, and, if relevant, multiplicity of analyses | 11 |
| Generalisability | 21 | Generalisability (external validity, applicability) of the trial findings | 11 |
| Interpretation | 22 | Interpretation consistent with results, balancing benefits and harms, and considering other relevant evidence | 10-11 |
| **Other information** | | |  |
| Registration | 23 | Registration number and name of trial registry | 4 |
| Protocol | 24 | Where the full trial protocol can be accessed, if available | n/a |
| Funding | 25 | Sources of funding and other support (such as supply of drugs), role of funders | 13 |

*We strongly recommend reading this statement in conjunction with the CONSORT 2010 Explanation and Elaboration for important clarifications on all the items. If relevant, we also recommend reading CONSORT extensions for cluster randomised trials, non-inferiority and equivalence trials, non-pharmacological treatments, herbal interventions, and pragmatic trials. Additional extensions are forthcoming: for those and for up to date references relevant to this checklist, see [www.consort-statement.org](http://www.consort-statement.org).
